# Concurrent mapping of Electrical, Chemical, and Functional Neuroactivity

**DOI:** 10.1101/2024.12.06.24317948

**Authors:** Zhifang Wu, Yameng Gu, Jianbo Cao, Sha Liu, Lingzhi Hu, Yang Li, Meng Liang, Hao Song, Shuai Yang, Jianmin Yuan, Jun Xie, Jun Xu, Xiaomeng Li, Rong Zheng, Hua Wei, Haiyan Liu, Hongliang Wang, Xinzhong Hao, Yujie Hu, Zhifen Liu, Lijun Yang, Yanbo Zhang, Xishan Sun, Ruokun Xu, Shizhong Lian, Quan Zhu, Weiwei Xie, Chunxia Xie, Di Zhang, Sijin Li

**Affiliations:** Department of Nuclear Medicine, First Hospital of Shanxi Medical University, Taiyuan, 030001, Shanxi, China; Shanxi Key Laboratory of Molecular Imaging, Shanxi Medical University, Taiyuan, 030001, Shanxi, China; Collaborative Innovation Center for Molecular Imaging of Precision Medicine, Shanxi Medical University, Taiyuan, 030001, Shanxi, China; United Imaging Healthcare, 201807, Shanghai, China; Department of Psychiatry, First Hospital of Shanxi Medical University, Taiyuan, 030001, Shanxi, China; Department of Neurology, First Hospital of Shanxi Medical University, Taiyuan, 030001, Shanxi, China; Department of Biochemistry and Molecular Biology, Shanxi Medical University, Taiyuan, 030001, China; Department of Hepatobiliary Surgery, First Hospital of Shanxi Medical University, Taiyuan, 030001, Shanxi, China; Department of Pharmacology, Shanxi Medical University, Taiyuan, 030001, Shanxi, China; Department of Health Statistics, School of Public Health, Shanxi Medical University, Taiyuan, 030001, Shanxi, China; Department of Neurosurgery, First Hospital of Shanxi Medical University, Taiyuan, 030001, Shanxi, China

## Abstract

Understanding the complex mechanism of human brain function requires innovative approaches that capture the intricate interplay of electrical, chemical, and hemodynamic activities. We developed the PMEEN system, named for its concurrent integration of **P**ET, **M**RI, **E**EG, **E**ye Tracking, and f**N**IRS modalities by successfully addressing the electromagnetic and gamma ray interference among these modalities and incorporation of centralized clock control to allow simultaneous spatial registration and temporal synchronization. Here we show that PMEEN enables the concurrent acquisition of metabolic, structural, electrical, hemodynamic, and behavioral data, providing a comprehensive view of brain function that was previously inaccessible. By successfully validating the system in healthy volunteers and clinical cases involving patients with major depressive disorder, Alzheimer’s disease, and epilepsy, we demonstrate its potential for advancing both research and clinical diagnostics. These findings represent a significant technological step forward in neuroimaging, allowing for a holistic understanding of the neural mechanisms underlying cognition and neurological disorders, and setting the stage for future breakthroughs in brain research and therapy.

**Summary:** We introduce the PMEEN system, a groundbreaking multi-modal neuroimaging platform integrating PET, MRI, EEG, Eye tracking, and fNIRS. This system allows concurrent acquisition of metabolic, structural, electrical, hemodynamic, and behavioral data, providing unprecedented insights into brain function. Through advanced electromagnetic compatibility design and synchronized data acquisition, PMEEN overcomes the technical challenges of integrating these modalities. Validated in phantom, healthy volunteers and patients with major depressive disorder, Alzheimer’s disease, and epilepsy, PMEEN has demonstrated its capability for capturing comprehensive neural dynamics. This system paves the way for new research opportunities and clinical applications in neurological and psychiatric diagnostics.

## Introduction

Interrogating brain function at rest and in response to external visual stimuli through non-invasive methods is crucial not only for basic neuroscience research but also for diagnosing and treating a wide spectrum of neurological and psychiatric disorders, such as Alzheimer’s disease, epilepsy, and major depressive disorder^1,2^. Visual processing serves as an exemplary model for studying neural dynamics, as it involves rapid signal transmission from the retina through the optic nerve to the visual cortex, engaging multiple neural pathways and processing centers^3,4^. This process encompasses a complex cascade of electrophysiological events, neurotransmitter release, and hemodynamic responses occurring within microsecond, highlighting the need for imaging modalities that can capture these fast-paced interactions with high spatial and temporal resolution. Understanding the intricate interplay of electrical, chemical, and functional activities in the human brain is fundamental to unraveling the complexities of cognition, perception, and consciousness.

Recent developments in non-invasive brain imaging techniques have significantly advanced our understanding of neural mechanisms. Magnetic Resonance Imaging (MRI), especially ultra-high-field systems operating at 3-7 Tesla and above, has improved spatial resolution and signal-to-noise ratio, enabling detailed structural and functional mapping of the brain’s microarchitecture^5,6^. High-resolution Positron Emission Tomography (PET) imaging, using advanced detector technologies and reconstruction algorithms, has enhanced our ability to visualize metabolic and molecular processes with greater precision^7,8^. Concurrently, real-time electroencephalogram (EEG) provides microsecond-scale temporal resolution of neuronal electrical activity, capturing oscillatory dynamics and event-related potentials essential for understanding cognitive processes^9^. Functional near-infrared spectroscopy (fNIRS) offers a portable and non-invasive means to monitor cerebral hemodynamics and oxygenation, complementing other imaging modalities by providing insights into neurovascular coupling^10,11^. Eye-tracking technology, by capturing gaze patterns and pupillary responses, serves as a window into attentional processes and autonomic nervous system activity^12,13^.

Despite these advancements, capturing the transient and multidimensional brain resting state and responses of the brain to visual stimuli remain challenging. Traditional imaging systems often operate in isolation or combine only two modalities, limiting the ability to comprehensively assess the simultaneous electrical, metabolic, and hemodynamic changes occurring during neural activity^14^. Integrating multiple imaging modalities poses significant engineering and physics challenges due to factors such as electromagnetic interference and gamma ray attenuation. Specifically, MRI systems with strong magnetic fields, high power radio-frequency emission and rapidly switching gradients, can distort signals from other modalities like EEG and PET. The interaction of gamma photons emitted during PET imaging with external materials in the field of view can introduce artifacts and degrade image quality^15^. Furthermore, achieving precise spatial and temporal synchronization across different modalities is also prohibitively difficult in its engineering implementation, and variations in data acquisition rates and timing accuracy can lead to misalignment and hinder the fusion of multimodal data^16,17^.

To overcome these boundaries, we have developed a novel multimodal imaging system, PMEEN (**P**ET, **M**RI, **E**EG, **E**ye tracking, and functional **N**ear-Infrared Spectroscopy), designed to achieve millimeter spatial resolution and microsecond temporal synchronization. As part of the China Brain Project^18^, PMEEN represents a fully integrated platform that synchronizes multiple imaging modalities through advanced electromagnetic compatibility (EMC) design, centralized electronic synchronization, and unified control software. The system incorporates innovative electromagnetic shielding techniques, noise reduction strategies, and hardware innovations to mitigate interference among modalities. Additionally, we have developed post-processing algorithms and tools to accurately fuse multimodal data in three dimensions, enabling detailed mapping of neural activity across different spatial and temporal scales. The performance of PMEEN is demonstrated through pilot studies involving healthy volunteers and patients with major depressive disorder, epilepsy, and Alzheimer’s disease, highlighting its potential for advancing neuroscience research and improving clinical investigation of neurological disease.

## Results

### System Design

The PMEEN system (Fig. 1) integrates **P**ET, **M**RI, **E**EG, **E**ye tracking, and functional **N**ear-Infrared Spectroscopy (fNIRS) technologies into one imaging bore (Fig. 2). The connections and communication pathways between various components are shown in a detailed schematic of the PMEEN system architecture (Fig. S1). At its core, the system captures electrical, chemical and functional signals of neurophysiological signals from multiple modalities through a simultaneous data acquisition software (Fig. S2). PET detects metabolic activity via gamma ray photons, while MRI provides high-resolution structural and functional brain images using a superconductive magnet. EEG measures electrical activity from the brain through sensors placed on the scalp, and fNIRS monitors hemodynamic changes by using light detectors and sources to assess blood oxygenation. An eye tracking camera captures visual attention and eye movement during visual stimuli presentations. Central to the system is a precise 0.1 microsecond synchronization clock, which ensures temporal alignment across all modalities, enabling real-time data acquisition. The system also incorporates electromagnetic compatibility (EMC) filtering and shielding to tackle interference among different modalities, along with gamma ray transparency and attenuation correction for accurate PET imaging. The PMEEN system generates multimodal data, including metabolic, structural, electrical, hemodynamic brain maps and eye movement, providing a comprehensive view of brain function and behavior. The data collected by the PMEEN system was validated through both phantom testing and human task experiment.

**Figure 1.**
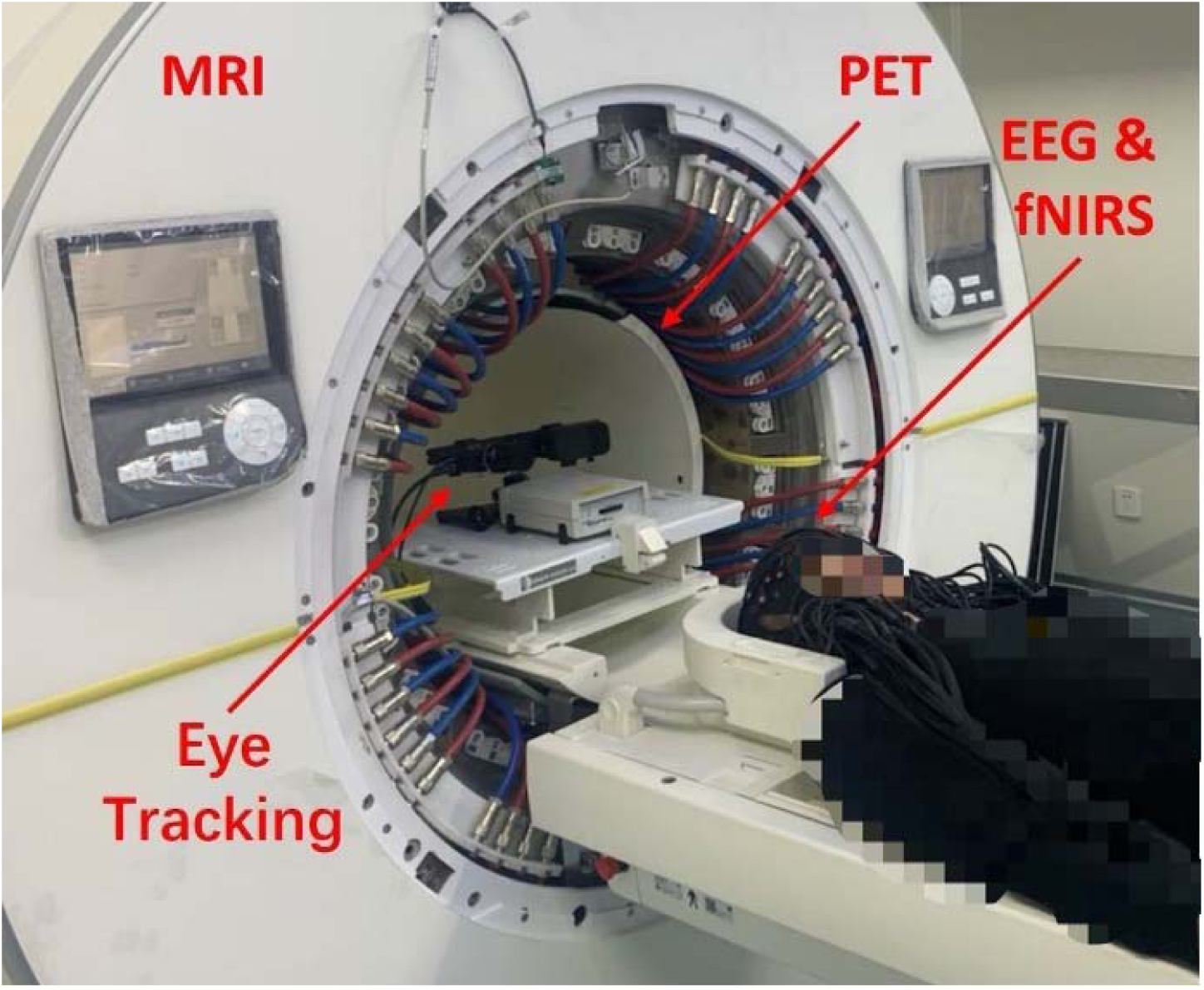
A picture of the PMEEN system without cover.

**Figure 2.**
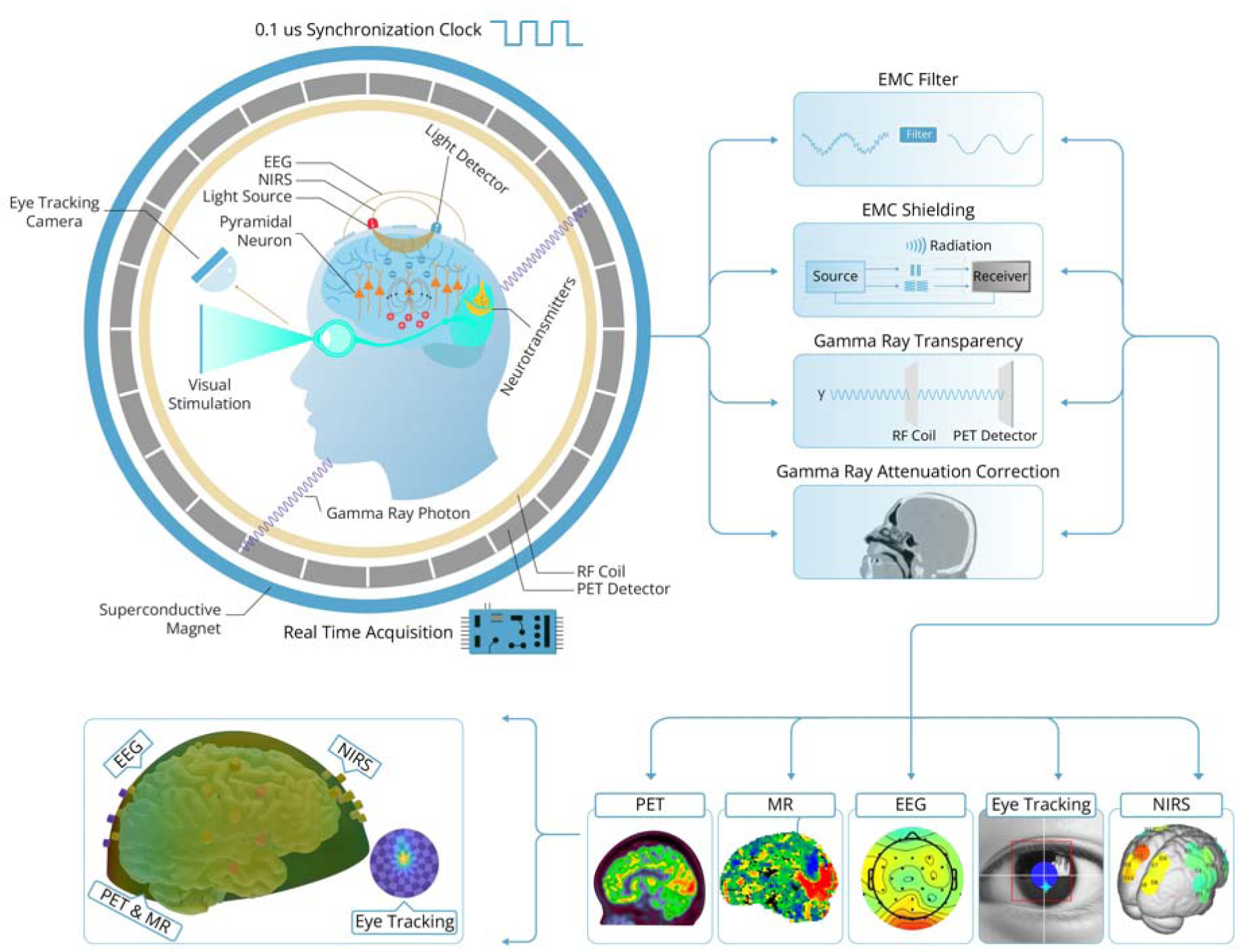
PMEEN System diagram. Real time acquisition of all imaging modalities is achieved by a centralized clock control. Electromagnetic Compatibility (EMC) of all imaging modalities is achieved by filtering and shielding design. To allow maximal quality for PET imaging, all material in FOV was either redesigned with gamma ray transparent material or corrected for its gramma tray attenuation. Data from all imaging modalities are registered and projected into the identical 3D space via a dedicated post-processing software for visualization and further quantitative process.

### System Validation

The MR image quality produced by the PMEEN system successfully met the standards of the American College of Radiology (ACR) MRI accreditation (Fig. S3 and Table S1). The MR ACR results were obtained under two conditions: with MR-only imaging and with the presence of an active PET acquisition, EEG electrodes, Eye-tracking mirror, fNIRS optodes and optic fiber (EEN) positioned on the phantom. In terms of uniformity, a slight decrease was observed when the PET acquisition was on and EEN components were included, with values of 92% for MR-only and 91% for the combined setup. Spatial resolution remained consistent at 1 mm for both conditions, while slice thickness accuracy exhibited minimal difference, with values of 0.12 mm and 0.17 mm, respectively. Percent signal ghosting increased from 0.10% in MR-only to 0.32% with the PET acquisition and EEN components. The signal-to-noise ratio was comparable between both setups, with values of 1555.4 for MR-only and 1553.4 for the condition with PET acquisition and EEN components. Despite these differences, both setups met ACR criteria, concluding a passing performance.

The PET image quality collected from the PMEEN system, assessed using the NEMA standard image quality (IQ) phantom is shown in Fig. S4 and Table S2. The NEMA IQ results were obtained with the MR sequence (BOLD EPI) active, and the EEN components positioned on the phantom. The hot contrast values for six spheres were all within the acceptable ranges, with values increasing from 66.22% for the smallest sphere to 90.75% for the largest. The hot contrast values exceeded the minimum acceptable thresholds for all sphere sizes, indicating good image contrast performance. Background variability, which decreased from 7.24% for the smallest sphere to 3.60% for the largest, also remained within the acceptable limits for each sphere size. The lung residual error was 3.7%, well below the acceptable threshold of 10%. Additionally, contrast recovery decreases as background variability increases for the PET IQ phantom under two conditions: with the phantom alone and with the phantom fitted with EEN components (Fig. S5). A uniform cylindrical phantom was used to evaluate the PET SUV changes induced by the EEN components in the PMEEN system. After applying attenuation correction for the EEN components, the PET SUV changes induced by the EEN components were found to be less than 5% (Fig. S6).

In addition, the quality of EEG, eye tracking and fNIRS imaging were validated using volunteer scans. Specifically, a visual checkerboard task with protocol specified in Fig. S7. BOLD-fMRI analysis revealed significant activation during task-on conditions, primarily in occipital and cingulate areas (top, Fig. 3A), including the occipital pole, lingual gyrus, lateral occipital cortex of inferior division, occipital fusiform gyrus, intracalcarine cortex, cerebellum, cingulate gyrus and paracingulate gyrus. Static FDG-PET data (bottom, Fig. 3A) showed activation pattern primarily in occipital area, with activity observed in the occipital pole, intracalcarine cortex, precuneous cortex, and lingual gyrus. Additionally, we observed an increase in oxygenated hemoglobin (HbO) during task-on conditions in channels located over the occipital regions (Fig. 3B), predominantly in the left visual cortex, while deoxygenated hemoglobin (HbR) did not show a similar increase. Compared to task-off conditions, neither oxygenated nor deoxygenated hemoglobin exhibited significant activation. The mean EEG spectrum, averaged across trials and channels located over the occipital cortex, showed peaks at 8 Hz, 16 Hz, and 24 Hz during task-on conditions (Fig. 3D), compared to task-off conditions, The frequencies at 8 Hz, 16 Hz, and 24 Hz correspond to the checkerboard’s driving frequency and its harmonics, respectively. Activation was only observed in the occipital region when comparing signal power difference at these peak frequencies between task-on and task-off conditions (Fig. 3D). A peak at 17 Hz, present in both task-on and task-off conditions, was attributed to MR helium pump noise. Pupil size significantly decreased (p < 0.001) during the task-on condition compared to the task-off condition (Fig. 3C), and eye-tracking heat maps indicated that participants focused on the center of the checkerboard during the task (Fig. 3C).

**Figure 3.**
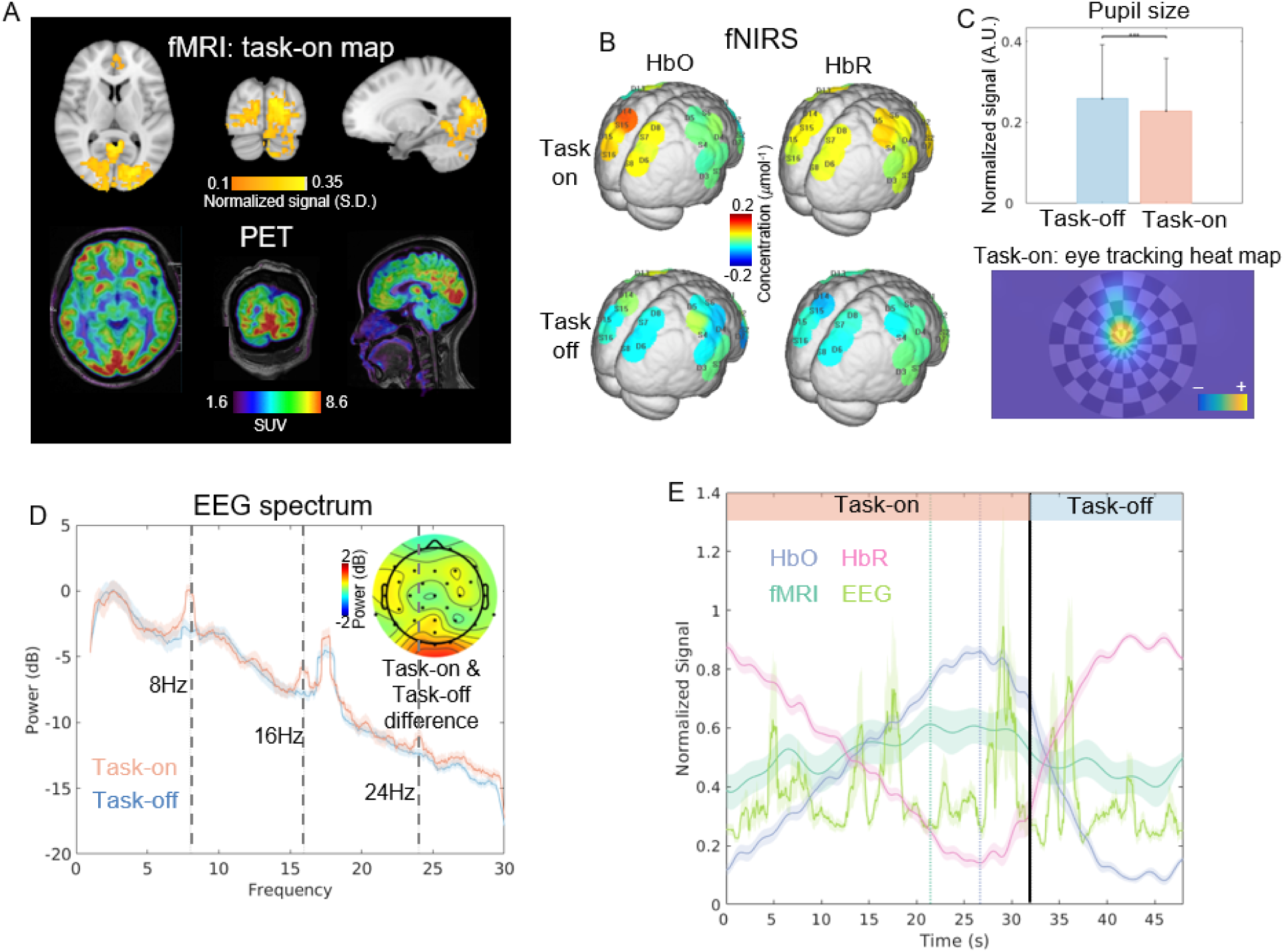
Results from the PMEEN system during the visual checkerboard task. A. Top: fMRI activation map during the checkerboard task-on conditions. Bottom: Static PET image for the entire task window. B. Mean HbO and HbR levels during task-on and task-off conditions, derived by averaging across time points within each condition and across trials. C. Top: Mean pupil size during task-on and task-off conditions, derived by averaging across time points within each condition and across trials. Bottom: Eye-tracking heat map for task-on conditions. D. Mean EEG spectrum during task-on and task-off conditions, derived by averaged across three occipital channels and across trials. The signal power difference at peak frequencies (8Hz, 16Hz, and 24Hz) between task-on and task-off conditions was also shown. E. Mean BOLD fMRI, HbO, and HbR time courses during task-on and task-off conditions, averaged over trials. The EEG time course was calculated by averaging the spectrogram at 8 Hz, 16 Hz, and 24 Hz across three occipital channels.

To illustrate the sequential activation across brain imaging modalities, we plotted the time courses of mean EEG power at the checkerboard’s driving frequency and its harmonics over three channels located at the occipital cortex, BOLD-fMRI activation in the occipital cortex, and HbO and HbR levels during both task-on and task-off conditions (Fig. 3E). The EEG power showed peaks at approximately 5 seconds, followed by BOLD-fMRI and HbO peaks at 21.44 and 26.66 seconds respectively. To assess the signal quality of the five simultaneously collected PMEEN imaging modalities, we collected the EEG, fNIRS and eye tracking signals during the same visual checkerboard task outside the PMEEN scanner and collected only BOLD fMRI during task without the other modalities. The same brain activation patterns were observed (Fig. S8).

### Clinical Application

We then applied the PMEEN system to image patients with three different neurological disorders, with one patient per disorder, each paired with the same healthy control. This was done with the goal of paving the way for future potential clinical applications of the PMEEN system.

The static FDG-PET imaging of the patient with major depressive disorder (MDD) revealed hypometabolism in the left frontal lobe, by comparing it with a group of health controls in the PET database (NeuroQ database) (Fig. 4A). To explore this further with other modalities, we computed a BOLD-fMRI correlation map using the left frontal lobe as the seed region. The patient with MDD exhibited positive correlations in the insular cortex, frontal orbital cortex, frontal medial cortex, cingulate gyrus, planum temporale, and subcallosal cortex, and negative correlations in the central opercular cortex, precentral gyrus, lateral occipital cortex (superior division), superior parietal lobule, precentral gyrus, and insular cortex. In contrast, the healthy control demonstrated broader and stronger positive correlations in the middle frontal gyrus, superior frontal gyrus, paracingulate gyrus, frontal medial cortex, precuneus cortex, temporal pole, frontal pole, left accumbens, and superior temporal gyrus (anterior division), and negative correlations in the precentral gyrus, lateral occipital cortex (superior division), superior parietal lobule, angular gyrus, supramarginal gyrus (anterior division), frontal pole, and cerebellum (Fig. 4B). The EEG spectrogram averaged over channels within the left frontal lobe showed that, in the healthy control, there was a prominent 12 Hz frequency band across the entire scan and some low-frequency activity around 3 Hz (Fig. 4C). In contrast, the MDD patient demonstrated an 8 Hz frequency band almost throughout the scan, with occasional low 2 Hz frequency activity (Fig. 4C). The EEG correlation map, using each of the channel within the left frontal lobe as the seed, indicated that the MDD patient had increased connectivity (values >0.5) with other channels primarily in the delta (0.5–4 Hz) and theta (4–8 Hz) bands (Fig. 4C). In comparison, the healthy control had higher connectivity (values >0.5) in the alpha (8–12 Hz) and beta (12–30 Hz) bands (Fig. 4C). The fNIRS HbO-based functional connectivity matrix indicated that the MDD patient had increased connectivity within the parietal and occipital cortices, but reduced connectivity in the frontal lobes, relative to the healthy control (Fig. 4D). Similarly, using each of the fNIRS channel within the left frontal lobe as a seed, the fNIRS correlation map in the MDD patient showed decreased connectivity (values>0.5) compared to the healthy control (Fig. 4D). Finally, the mean pupil size of the MDD patient during scanning was significantly reduced (p < 0.001) compared to that of the healthy control (Fig. 4E).

**Figure 4.**
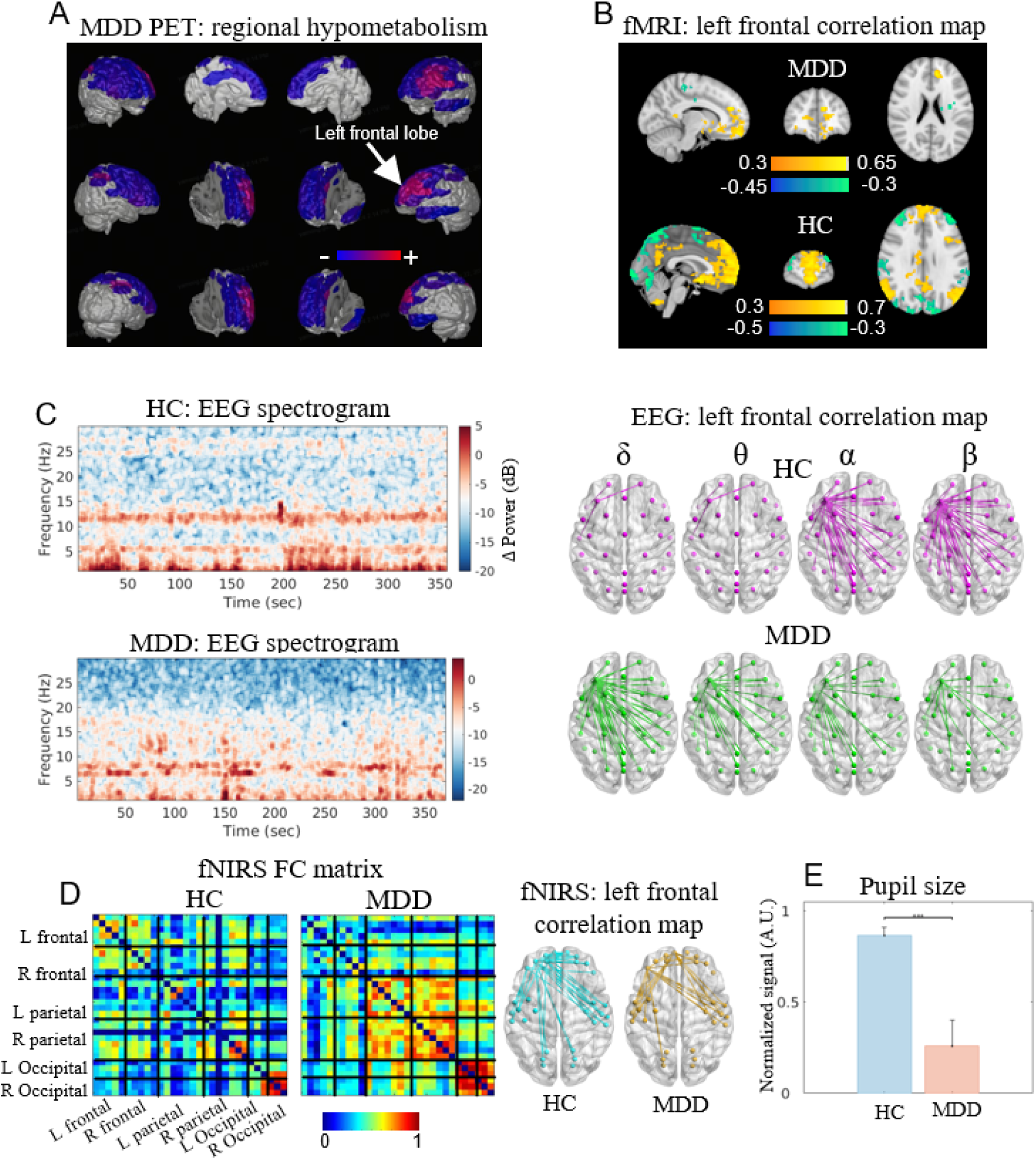
Results from the PMEEN system for the MDD patient during the resting-state. A. Brain regions with hypometabolism identified by comparing FDG-PET images of the MDD patient with those of the healthy control group (NeuroQ database). B. Correlation map using the left frontal lobe as the seed region in both the healthy control and the MDD patient. C. Left: Mean EEG spectrogram over channels located in the left frontal lobe for both the healthy control and the MDD patient. Right: Correlation map using each of the EEG channels in the left frontal lobe as the seed region for both the healthy control and the MDD patient. D. Left: Functional connectivity matrix calculated from fNIRS HbO signals in the healthy control and the MDD patient. Right: Correlation map of HbO signals using each of the fNIRS channels in the left frontal lobe as the seed region for both the healthy control and the MDD patient. E. Mean pupil size averaged over the scan for both the healthy control and the MDD patient.

Static FDG-PET imaging of the Alzheimer’s disease (AD) patient revealed hypometabolism in the left and right frontal lobes, left and right parietal lobes, and the cingulate cortex, comparing with a group of healthy controls (NeuroQ database) (Fig. 5A). To further explore these findings across other modalities, we generated BOLD-fMRI (Fig. 5B), EEG (Fig. 5C, right), and fNIRS (Fig. 5D, right) correlation maps using each brain region showing hypometabolism as a seed region. In the left frontal seed map, the AD patient showed positive correlations in the superior frontal gyrus, cingulate gyrus, and left thalamus, with negative correlations in the lateral occipital cortex and superior parietal lobule. Note that only a subset of brain regions is listed here due to space constraints; the full list can be found in the Supplementary Results. The healthy control displayed broader positive correlations in the middle and superior frontal gyrus, and negative correlations in the precentral gyrus. In the right frontal seed map, the AD patient had positive correlations in the superior frontal and angular gyri, while the healthy control showed positive correlations in the precentral gyrus and left thalamus, and negative correlations in the insular cortex and posterior cingulate gyrus. For the left parietal seed map, the AD patient showed positive correlations in the postcentral gyrus and supplementary motor cortex, and negative correlations in the precuneus and lingual gyrus. The healthy control had positive correlations in the superior frontal gyrus and superior parietal lobule, with negative correlations in the frontal orbital cortex and left thalamus. In the right parietal seed map, the AD patient had positive correlations in the precentral gyrus and superior parietal lobule, and negative correlations in the cuneal and lingual gyrus. The healthy control showed positive correlations in the superior parietal lobule and postcentral gyrus, with negative correlations in the posterior cingulate and occipital pole. In the cingulate seed map, the AD patient had positive correlations in the supplementary motor cortex and superior frontal gyrus, and negative correlations in the lateral occipital cortex, while the healthy control showed positive correlations in the superior and middle frontal gyri, and negative correlations in the lateral occipital cortex.

**Figure 5.**
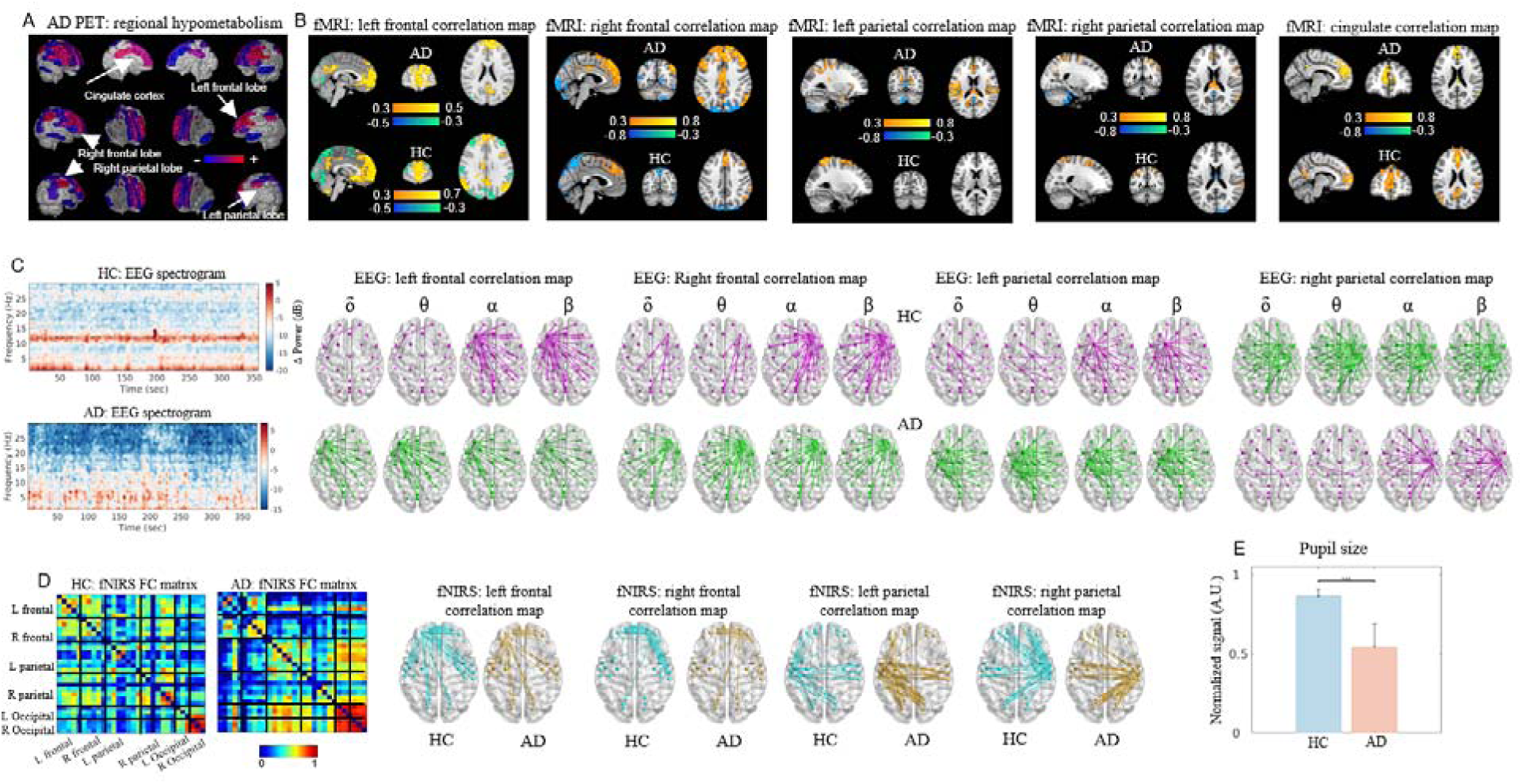
Results from the PMEEN system for the AD patient during the resting-state. A. Brain regions with hypometabolism identified by comparing FDG-PET images of the AD patient with those of the healthy control group (NeuroQ database). B. Correlation maps using the left frontal lobe, right frontal lobe, left parietal lobe, right parietal lobe, and cingulate cortex as seed regions respectively in both the healthy control and AD patient, respectively. C. Left: Mean EEG spectrogram over channels located in the brain regions of hypometabolism, including the left frontal lobe, right frontal lobe, left parietal lobe, right parietal lobe, and cingulate cortex for both the healthy control and AD patient. Right: Correlation maps using EEG channels in the brain regions of hypometabolism in both the healthy control and AD patient. D. Left: Functional connectivity matrix calculated from HbO signals collected by fNIRS in both the healthy control and AD patient. Right: Correlation maps of HbO signals using fNIRS channels in the left frontal lobe, right frontal lobe, left parietal lobe, and right parietal lobe as seed regions respectively in both the healthy control and AD patient. E. Mean pupil size averaged over the scan for both the healthy control and AD patient.

The EEG spectrogram averaged over hypometabolic regions showed that the healthy control had a prominent 12 Hz band and low-frequency activity around 1 Hz, while the AD patient displayed a 3–7 Hz band (Fig. 5C, left). The EEG correlation map with frontal lobe seeds indicated increased delta and theta connectivity in the AD patient and stronger alpha and beta connectivity in the control (Fig. 5C, right). The left parietal lobe map showed increased connectivity across all frequency bands in the AD patient, while the right parietal lobe map showed decreased connectivity (Fig. 5C, right). The cingulate cortex was excluded due to EEG’s cortical focus. fNIRS HbO connectivity maps revealed decreased connectivity in the AD patient’s frontal and right parietal lobes, but increased connectivity in the left parietal and occipital regions (Fig. 5D, left). Frontal and right parietal seed maps showed reduced connectivity, whereas the left parietal seed map showed increased connectivity compared to the control (Fig. 5D, right). Lastly, the AD patient’s pupil size during the scan was significantly smaller (p < 0.001) compared to that of the healthy control (Fig. 5E).

Further clinical evaluation of the patient can be found in supplementary information.

The periventricular nodular heterotopia (PVNH) in the epilepsy patient was visible in both T1-weighted and static FDG-PET images (Fig. 6A). Static FDG-PET revealed hypometabolism in the left frontal lobe compared to healthy controls (NeuroQ database) (Fig. 6B). A BOLD-fMRI correlation map using the left frontal lobe as the seed region showed positive correlations in the central opercular and insular cortices, and negative correlations in the superior frontal gyrus (Fig. 6C). The healthy control had stronger positive correlations in the middle and superior frontal gyri, and negative correlations in the precentral gyrus and superior parietal lobule (Fig. 6C). The fNIRS HbO-based connectivity matrix showed increased connectivity in the right parietal lobe of the epilepsy patient but decreased connectivity elsewhere (Fig. 6D). The fNIRS HbO correlation map also showed reduced connectivity in the left frontal lobe of the epilepsy patient compared to the control (Fig. 6D). Clinical reports indicated that the patient’s seizures originated from EEG channels F7, F8, T7, and T8, so we analyzed the EEG spectrogram averaged across these channels. The epilepsy patient showed a consistent 10 Hz band, while the healthy control exhibited a 12 Hz band, 1 Hz band, and occasional 3 Hz activity (Fig. 6E). The EEG correlation map using F7 and F8 as seed regions revealed increased connectivity in the delta and theta bands and decreased connectivity in the alpha and beta bands in the epilepsy patient compared to the control (Fig. 6E). Using T7, the patient showed increased connectivity in the delta, theta, and alpha bands but decreased beta connectivity, while T8 showed increased connectivity across all bands (Fig. 6E). The patient’s pupil size was significantly smaller than the healthy control’s (p < 0.001) (Fig. 6F).

**Figure 6.**
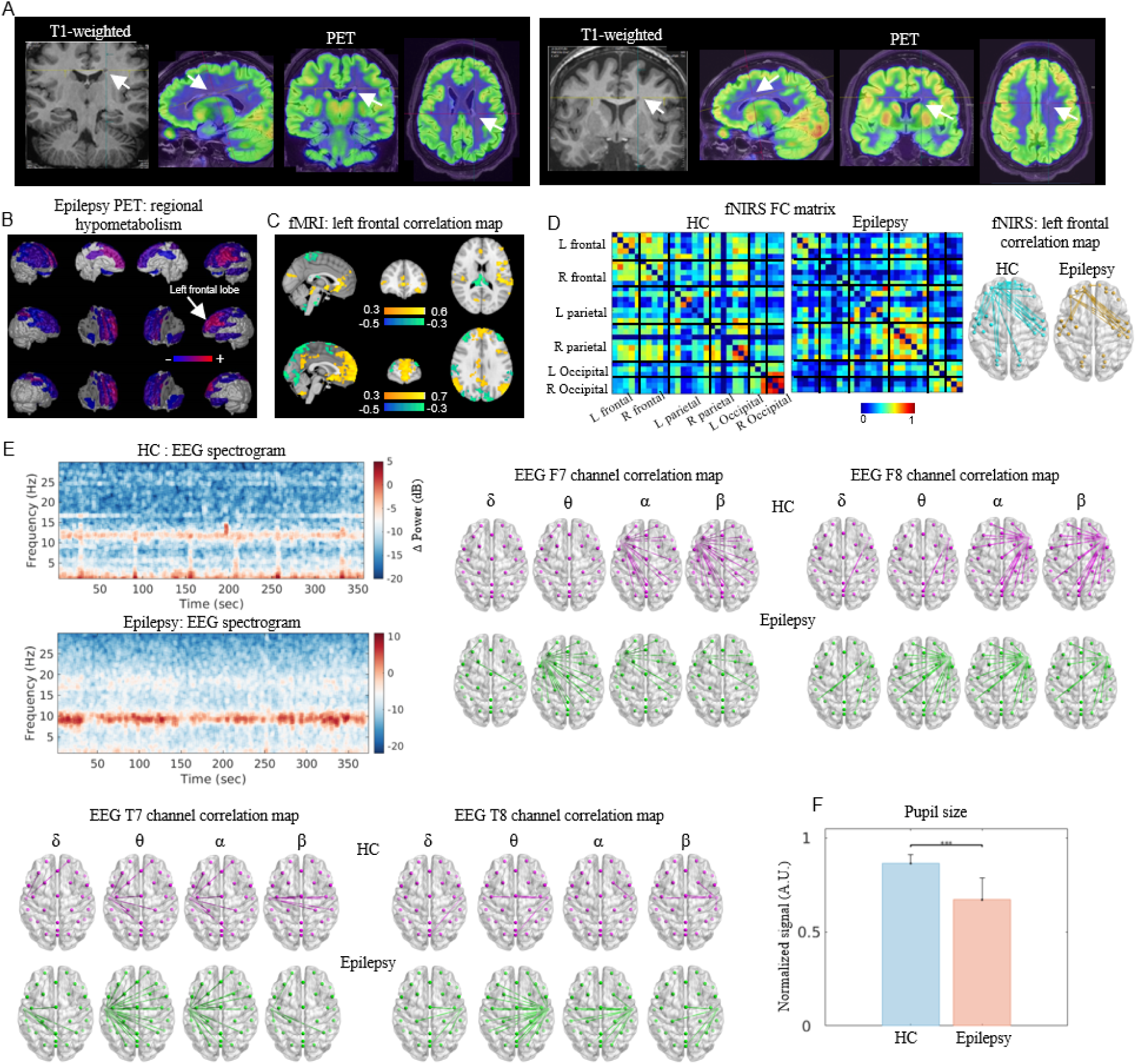
Results from the PMEEN system for the epilepsy patient during the resting-state. A. T1-weighted image and static FDG-PET image detect periventricular nodular heterotopia in the epilepsy patient, indicated by the white arrow. B. The brain regions with hypometabolism identified by comparing FDG-PET images of the epilepsy patient with those of the healthy control group (NeuroQ database). C. Correlation maps using the left frontal lobe as seed regions in both the healthy control and epilepsy patient, respectively. D. Left: Functional connectivity matrix calculated from HbO signals collected by fNIRS in both the healthy control and epilepsy patient. Right: Correlation maps of HbO signals using fNIRS channels in the left frontal lobe as seed regions in both the healthy control and epilepsy patient. E. Left: Mean EEG spectrogram over the F7, F8, T7, T8 channels for both the healthy control and epilepsy patient respectively. Right: Correlation maps using F7, F8, T7, T8 channels as seed regions in both the healthy control and epilepsy patient respectively. E. Mean pupil size averaged over the scan for both the healthy control and epilepsy patient.

Further clinical evaluation of the patient can be found in supplementary information.

## Discussion

We have developed and validated the PMEEN system, the first of its kind to simultaneously acquire data from PET, MRI, EEG, eye-tracking, and fNIRS modalities. This integrated platform enables comprehensive assessment of brain function by capturing electrical, metabolic, hemodynamic, and behavioral responses with high spatial and temporal resolution. The implementation of advanced EMC shielding, and gamma-ray correction technologies ensures minimal interference among modalities, allowing each to achieve optimal performance concurrently. Our pilot studies demonstrate the system’s capability to capture immediate neural responses to visual stimulation, revealing intricate patterns of neural activation and connectivity that were previously inaccessible with single-modality or sequential imaging approaches.

The integration of multiple imaging modalities within a single system addresses a longstanding challenge in neuroscience—the ability to correlate fast electrophysiological events with slower hemodynamic and metabolic changes in real time. EEG provides direct measurements of neuronal electrical activity with microsecond temporal resolution but lacks spatial specificity, while fMRI offers high spatial resolution but with a temporal lag due to the hemodynamic response^4^. PET imaging adds a molecular dimension, enabling the visualization of metabolic processes and neurotransmitter systems, but traditionally suffers from low temporal resolution^19^. By synchronizing these modalities, PMEEN bridges the gap between temporal and spatial scales, facilitating a more holistic understanding of neural dynamics.

Overcoming the technical challenges associated with multimodal integration required several innovative solutions. The strong magnetic fields, fast-switching gradient and RF pulses of MRI can induce artifacts and distortions in EEG and PET signals. We addressed this by designing specialized EEG caps and fNIRS optodes using non-magnetic, MRI-compatible materials, and implementing active shielding techniques to protect sensitive electronics^20^. For PET imaging, we developed correction algorithms to account for attenuation and scatter caused by the EEG and fNIRS equipment within the field of view, using UTE MRI sequences for precise attenuation maps^21^. A centralized clock distribution system ensures sub-microsecond synchronization across all modalities, critical for accurate temporal alignment and data fusion^17^. The unified control software streamlines data acquisition and processing, reducing the potential for human error and enhancing reproducibility.

The preliminary applications of PMEEN in clinical populations highlight its potential impact on neurological and psychiatric diagnostics. In the case of major depressive disorder (MDD), the system detected hypometabolism in the left frontal cortex through FDG-PET, consistent with previous findings implicating frontal lobe dysfunction in mood regulation^22^. Concurrent EEG and fNIRS measurements revealed altered electrophysiological patterns and hemodynamic responses, suggesting disrupted neurovascular coupling in MDD^23^. Eye-tracking data showed difficulties in maintaining sustained attention, correlating with clinical symptoms of cognitive impairment in major depressive disorder^24^.

In Alzheimer’s disease, PMEEN identified characteristic patterns of hypometabolism in the posterior cingulate cortex and temporoparietal regions, areas known to be affected early in the disease process^25^. Functional connectivity analyses from fMRI and fNIRS data revealed network disruptions consistent with default mode network alterations observed in AD^26^. EEG data showed slowing of cortical rhythms, and eye-tracking metrics indicated impaired oculomotor function, which may reflect attentional deficits and executive dysfunction^27,28^.

In epilepsy, high-resolution MRI detected subtle structural anomalies indicative of periventricular nodular heterotopia, a known cause of refractory epilepsy^29^. PET imaging revealed areas of hypometabolism that may correspond to epileptogenic foci. Although the EEG did not capture abnormal activity during the interictal period, the multimodal data provided valuable insights into the patient’s condition, potentially guiding surgical planning or therapeutic interventions.

The development of PMEEN holds significant promise for advancing our understanding of brain function and pathology. By enabling simultaneous, multimodal imaging, researchers can explore the relationships between neuronal activity, hemodynamics, metabolism, and behavior with unprecedented detail. This integrated approach is particularly valuable for studying complex neural phenomena such as neuroplasticity, functional connectivity, and the pathophysiology of neuropsychiatric disorders^30,31^. For example, in cognitive neuroscience, PMEEN can be used to investigate the neural correlates of consciousness, attention, and sensory processing by capturing rapid electrophysiological events alongside slower hemodynamic changes and metabolic alterations^32^. In clinical research, the system offers a powerful tool for early diagnosis, treatment monitoring, and the evaluation of therapeutic interventions across a range of neurological conditions.

Despite these advancements, several limitations must be addressed in future work. The current configuration of the PMEEN system limits EEG and fNIRS channels to 32 due to space constraints within the headcap. While the current setup is sufficient for many applications, higher-density arrays would improve spatial resolution and source localization accuracy^33^. Future designs should focus on miniaturizing sensors and optimizing cap ergonomics to accommodate more channels. Additionally, the use of FDG as the PET tracer provides valuable metabolic information but lacks specificity for neurotransmitter systems. Incorporating radioligands targeting specific neurotransmitter receptors, such as dopamine or serotonin, would enhance the system’s ability to investigate neurochemical alterations in psychiatric disorders^34^. Moreover, the use of checkerboard visual stimulation, while effective for system validation, represents a simplistic stimulus. Employing more complex, ecologically valid tasks would allow for the examination of neural responses under conditions that more closely resemble real-world experiences^35^. Finally, expanding the sample size and diversity of clinical populations is essential for validating the clinical utility of PMEEN. Larger studies with well-characterized cohorts would enable the identification of biomarker profiles and the development of predictive models for disease progression and treatment response^36^. Collaborations with multiple research centers could facilitate multicenter trials, enhancing the generalizability of findings and accelerating the translation of this technology into clinical practice.

In conclusion, the PMEEN system represents a major advancement in multimodal neuroimaging, offering unparalleled opportunities to investigate the complex interplay of electrical, chemical, and functional activities in the human brain. By capturing synchronized data across multiple modalities with microsecond and millimeter accuracy, PMEEN provides a comprehensive platform for advancing neuroscience research and improving clinical diagnostics, ultimately contributing to a deeper understanding of brain function in health and disease.

## Method

The present study was approved by the ethical committee of the First Hospital of Shanxi Medical University.

### System design specifications

A multimodal neuroimaging system (PMEEN) is designed according to the system connection diagram specified in Fig. S1 and a dedicated multi-modal signal collection software has been developed to simultaneously collect and process the signal of different imaging modalities (Fig. S2). An overview of how each modality complements the others was provided (Fig. 1).

The parameters and specifications of the PET and MR system are introduced below. The spatial resolutions in the axial, radial, and tangential directions are 2.72 mm, 2.86 mm, and 2.81 mm FWHM, respectively. The NECR peak is at 129.2 kcps with an activity concentration of 14.7 kBq mL−1. The scatter fraction at the NECR peak is 37.9%, and the maximum slice error below the NECR peak is 4.1%. Contrast recovery coefficients varied from 51.8% for the 10 mm hot sphere to 87.3% for the 37 mm cold sphere. The TOF resolution at an activity concentration of 5.3 kBq mL^−1^ is at 535 ps, while with a point source, the TOF resolution is 474 ps. The MRI system has a magnetic field strength of 3 Tesla, 96 radiofrequency (RF) channels, a peak gradient strength of 50 mT/m, and a slew rate of 200 T/m/s. A 24-channel phase-array head coil was used in this study.

EEG was built based on a 32-channel EEG system (BrainAmp, Brain Products GmpH, Germany). Electrode placement across the scalp adhered to the international10–20 System to achieve standardized coverage of cortical regions with AFz and FCz as the ground and reference electrodes, respectively (Fig. S9). An electrocardiogram (ECG) electrode was affixed to the participant’s back for concurrent cardiac signal recording. Clock synchronization between the EEG system and the MR system’s master clock was achieved using a phase locking device (Syncbox, Brain Products GmbH, Germany). The EEG amplifier and battery were positioned behind the scanner bore and connected to the outside recording software through a waveguide using a 30-meter-long fiber-optical cable. Before the scanning, impedances for all electrodes were reduced and maintained below 20 kΩ with a more stringent threshold below 10 kΩ set for the ground and reference electrodes to minimize noise and enhance signal clarity. The raw EEG data were captured at a high-fidelity sampling rate of 5000 Hz, processed through a band-pass filter set between 0.1 and 250 Hz to accurately record a wide range of neural frequencies.

Eye tracking was built based on the system which employed an infrared video camera (EyeLink 1000 Plus, SR Research) operating at a sampling frequency of 1000 Hz. The camera and battery were positioned behind the scanner bore and connected to the outside recording software through a waveguide using a 25-meter-long fiber-optical cable.

fNIRS was built based on three continuous waves (NirScan-500BFC, Danyang Huichuang Medical Equipment Co., Ltd., China) to measure concentration changes of oxygenation and de-oxygenation. The optical system comprised 16 light sources, operating at wavelengths of 730 and 850 nm, and 16 avalanche photodiodes detectors, forming 30 effective channels. The signals were sampled at a rate of 11 Hz. The precise positioning of the sources, detectors, and anchor points (located at Nz, Cz, Al, Ar, Iz referring to the standard international 10-20 system of the electrode placement) was measured by an electromagnetic 3D digitizer device (Patriot, Polhemus, USA) on one participant. The acquired coordinates were then transformed into Montreal Neurological Institute (MNI) coordinates and further projected to the MNI standard brain template using spatial registration approach in NirSpace (Danyang Huichuang Medical Equipment Co., Ltd., China). The MNI coordinates for each channel were listed in Table S1. The 8-meter-long fNIRS optical fibers were routed from the subjects’ heads, along the scanner bed, to the outside recording software through the waveguide.

Visual stimulation was presented binocularly using MR-compatible screen (Shenzhen Sinorad Medical Electronics Co., Ltd., HDMI, 1920 x 1080 pixels, refresh rate 60 Hz, color depth 16.7 million colors). A fiber optic cable connected the system to a control computer through the waveguide outside the scanner room.

To combine EEG electrodes and fNIRS optodes onto one head cap for PMEEN system, a unified cap that supports both EEG electrodes and NIRS optodes was designed by adding additional openings (M15) on the standard EEG 32Ch MR compatible BrainCap-MR (EASYCAP GmbH, Herrsching, Germany). The EEG electrodes were placed on the EasyCap according to the International 10-20 system. We then arranged the fNIRS optodes into the additional openings, optimizing their placement to minimize crosstalk and maximize spatial resolution. The average distance between the source and the detector is 3 cm (range 2.7-3.3 cm). The specific location of the fNIRS optodes can be found in Fig. S10.

### Solutions for Concurrent Multi-modal Imaging

The primary physical interactions among the various imaging modalities within PMEEN can be categorized into two main types: electromagnetic interference (EMI) and gamma-ray attenuation (Fig. 7). EMI is addressed through careful design of shielding materials that block electromagnetic waves from penetrating the receiver electronics of each imaging modality. In PMEEN, we utilized a combination of metal and carbon fiber shielding, along with precise grounding, to minimize interference. To further reduce residual interference, we implemented both electronic notch and band-pass filters, as well as software-based filtering, to eliminate crosstalk between the different modalities. Water cooling was introduced to minimize the heating generated from MR gradient pulsing.

**Figure 7.**
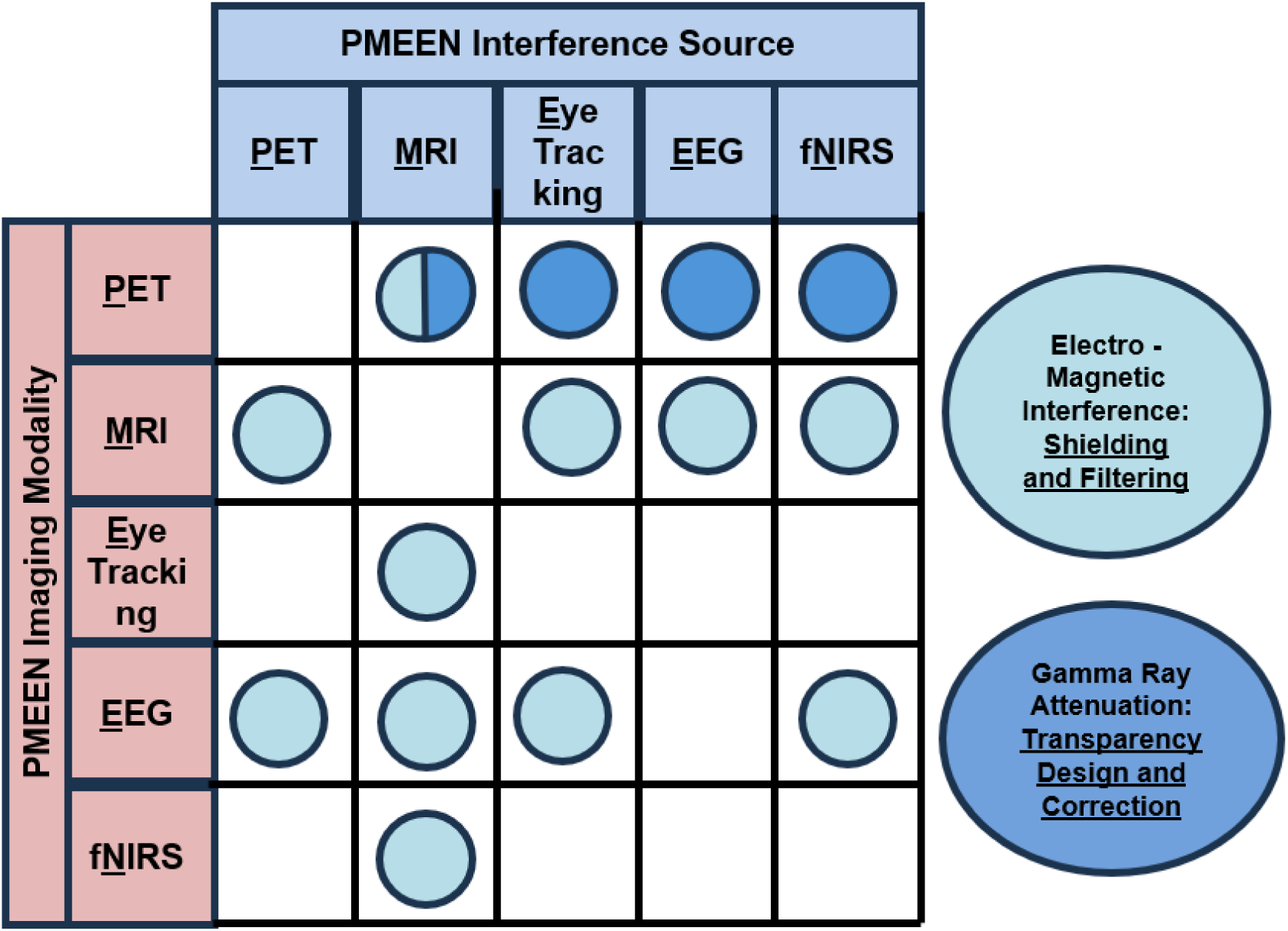
Mitigating interference among imaging modalities within PMEEN and engineering solutions for concurrent multi-modal imaging.

For gamma ray attenuation, we optimized the selection of hardware materials inside the PMEEN imaging bore to ensure maximum gamma-ray transparency for PET imaging. In cases where attenuation persisted, we employed an ultra-short echo time (UTE) MRI sequence to visualize attenuating hardware and applied a predefined linear attenuation coefficient (LAC) map to accurately correct for PET reconstruction artifacts. For attenuation arising from human brain, a water fat imaging sequence was employed to generate high resolution MRI image and an AI-based algorithm was used to translate T1 MRI into LAC map.

For further data preprocessing and data analysis, it is essential to achieve temporal synchronization in PMEEN system across PET, MR, EEG, eye tracking, and fNIRS systems. The PET images are automatically synchronized with MR signals via its control console. To temporally align the EEG, eye tracking, and fNIRS with the fMRI data, we use the MR gradient system trigger output. The MR gradient system trigger output generates a TTL (Transistor-Transistor Logic) pulse at the commencement of each functional MRI volume acquisition. Each TTL pulse is captured as a marker in the EEG, eye tracking, and fNIRS signals. Stimulus presentation was synchronized with the functional MRI acquisition by automatically starting the stimulus presentation as soon as the stimulus computer received the first functional MRI trigger signal.

### Phantom image quality evaluation

The impact of **E**EG electrodes, **E**ye-tracking mirrors, f**N**IRS optodes and optical fibers (EEN) components on the quality and quantification of the acquired MR or PET images collected from PMEEN system was examined.

The impact of EEN components on MR image was examined using the phantom following the American College of Radiology (ACR) MRI accreditation. After positioning the phantom at the center of the 24-channel head coil, images were acquired with a T1-weighted spin echo sequence (TR/TE = 500/30 ms, slice thickness=5mm, inter-slice gap = 5mm), and the scan was repeated twice to obtain the average results. Initially, images were captured with only the MR sequence active. Subsequently, the EEN components were placed on the phantom, and the PET sequence was activated to examine the multi-modal PMEEN system effect on MR images. The specific experimental setup can be found in Fig. S3. The results passed the ACR MRI accreditation (Table S1).

We utilized a NEMA image quality phantom (Data Spectrum Corp, NC, USA) to evaluate the impact of EEG electrodes, fNIRS optodes and optical fibers, and the eye tracking mirror on PET image quality collected from PMEEN system (Fig. S4). The NEMA image quality phantom was filled according to NEMA standards^37^ and positioned on the patient bed. The background was filled with F-18 FDG at an activity concentration of 11.15 kBq mL^−1^. The hot spheres contained an activity concentration four times that of the background, while the cold spheres were filled with water. Positioned centrally in the axial FOV, the phantom underwent a 10-minute scan with the MR system in idle mode under the phantom-only condition. To account for F-18 decay, the scanning, with MR pulsing (EPI sequence) activated and the EEG electrodes, fNIRS optodes and optical fibers, and eye-tracking mirror placed on the image quality phantom to mimic the conditions of PMEEN system, was extended to 13 minutes. The eye-tracking mirror was mounted on grey foam to maintain an 8 cm distance from the phantom, simulating the same distance between the human eye and the mirror as in human experiments. A CT-measured built-in phantom attenuation template was utilized for co-registration with the non-attenuation-corrected phantom image. Only the body transmission coil was within the FOV during scans. The standard contrast recovery curve versus background variability was plotted for all spheres under each condition (Fig. S5). The NEMA IQ results have passed the required standards (Table S2).

To assess the impact of EEN components on the SUV of PET images acquired from PMEEN system, we employed a 20-cm-diameter uniform cylindrical phantom with and without the presence of EEN components (Fig. S6). The uniform cylindrical phantom simulates the human brain during the simultaneous scanning of PMEEN system. The choice of this phantom was due to its sufficient uniform slices, which are ideal for assessing the effects on SUV. Additionally, we developed an algorithm for attenuation correction of the fNIRS optodes and optical fibers.

A uniform cylindrical phantom (radioactive concentration of 6.71 kBq mL^−1^ and a weight of 9.65 kg, 9650 mL) was filled and positioned on the patient bed within central axial FOV. The phantom was scanned for 10 minutes under the phantom-only condition. Since it was filled only once, the acquisition time was extended to 15 minutes with the EEN components placed on the phantom, to account for the decay of F-18 FDG. The eye-tracking mirror was mounted on the 24-channel head coil to simulate the human experiments. A built-in CT-derived attenuation correction template was employed for co-registering with the non-attenuation-corrected phantom images. No MR coils other than the body transmission coil were within the FOV. The mean SUV across each transverse slice was calculated using a circular region of interest covering approximately 90% of the phantom’s interior diameter. The middle slices with EEN components on the phantom were chosen to demonstrate the results.

To achieve the attenuation correction of the EEN components, we identified the EEN components using an ultra-short time echo (UTE) MR sequence, then assigned the mu values for attenuation correction. Given the high flexibility and variable placement of the EEN cables in each experiment, conventional fixed templates proved ineffective. Thus, we utilized the UTE to image the EEN components before each session. This automatic attenuation correction process for EEN components has been integrated into the PMEEN system, facilitating simultaneous scanning.

We conducted experiments to assess the effectiveness of attenuation correction for EEN components in the PMEEN system. A uniform cylindrical phantom was filled (radioactive concentration of 6.71 kBq mL^−1^ and a weight of 9.65 kg, 9650 mL) and positioned in the center of the 24-channel head coil. To account for the decay of F-18, the phantom under the phantom-only condition was initially scanned for 10 minutes. Additional scans, extending the acquisition time to 15 minutes, were performed with EEN components placed on the phantom and eye-tracking mirror mounted on the 24-channel head coil. The UTE MR images were then acquired to visualize the EEN components. Following the application of our automatic attenuation correction of the EEN components, the PET images demonstrated less than 5% SUV decrease across each slice of the phantom, comparable to those obtained from the phantom alone (Fig. S6).

### Multi-modal Imaging Visualization Platform

A platform was developed to integrate and visualize multi-modal imaging from the PMEEN system. The diagram in Fig. S11 represents how each modality undergoes specific preprocessing steps before being aligned and integrated for visualization. The processing of fMRI begins with skull stripping to remove non-brain tissue, followed by image segmentation to differentiate brain structures. Spatial smoothing is then applied, and the fMRI data is registered to MNI space by first aligning it with the T1-weighted images. Following these steps, nuisance regression is applied to remove confounding factors such as head motion. Finally, a temporal filter is applied to the time series fMRI data.

The other imaging modalities were first temporally aligned with the fMRI signal. In the PET modality, image reconstruction is the first step in processing the PET data. Since fNIRS optic fibers are used simultaneously, attenuation correction of the fNIRS optic fibers based on the UTE and WFI images is necessary to be included. Following this, skull stripping is performed to remove extraneous signals, and registration to MNI space ensures that the PET data aligns properly with fMRI. The fNIRS data undergoes several preprocessing steps to ensure data accuracy. First, spike artifacts—sudden, irregular disturbances in the data—are corrected. A band-pass filter is then applied. After preprocessing, the optical intensity data is converted into concentrations of HbO and Hb. Gradient artifacts from the MR scanner are removed from the EEG data, followed by the application of a low-pass filter and down-sampling. Next, pulse artifacts, often caused by the heartbeat, are eliminated. The data is then further cleaned by removing artifacts caused by muscle activity, eye blinks, or eye movements. The eye-tracking process begins with removing artifacts caused by eye blinks, followed by detrending the pupil size to eliminate slow drifts. Next, the envelope of the pupil size is calculated to smooth out fluctuations, and the data is normalized for comparability across sessions or subjects.

At the end of the pipeline, the processed EEG and fNIRS data were projected into MNI space for alignment with the fMRI and PET data. The PET and fMRI data were then mapped from volume space to surface space using HCP Workbench^38^. Subsequently, all five imaging modalities were resampled to match the same temporal resolution. For concurrent visualization, the PET and fMRI images were blended using a weighted ratio that sums to 1, allowing the ratio to be adjusted as needed to emphasize specific image features. Additionally, EEG electrodes and fNIRS optodes were projected onto the external brain surface based on their MNI coordinates, with their signal intensities displayed using distinct color maps. Pupil size and gaze position are displayed alongside other brain imaging modalities.

### Acquistion of Healthy Volunteer Data with Visual Stimulation

Two participants were recruited for the experiment: a 46 to 50-year-old female for the PMEEN system and a 21 to 25-year-old male for the EEN experiment. Both participants were screened for conditions like diabetes, neurological disorders, and MRI compatibility. Female participants were also screened for pregnancy. Before the scan, participants followed a high-protein, low-sugar diet and fasted for six hours. Visual checkerboard stimulation was used to verify the quality of multimodal data collected in the PMEEN system. The task involved alternating task-on and task-off periods to create contrast for FDG-PET, fMRI, fNIRS, and EEG. Eye-tracking data were also compared between conditions. FDG-PET, fMRI, EEG, fNIRS, and eye-tracking data were collected simultaneously using PMEEN system, with specific parameters for each modality. Data were preprocessed, aligned, and corrected for artifacts. fMRI, fNIRS, EEG, and eye-tracking data were analyzed to assess visual activation, pupil size, and gaze position between task-on and task-off conditions. The detailed method is available in the Supplementary Method.

### Acquisition of Patient Resting-State Data

One healthy female participant (41 to 45 years old) and three participants with specific conditions (a 36 to 40-year-old male with major depressive disorder, a 56 to 60-year-old female with Alzheimer’s disease, and a 26 to 30-year-old female with epilepsy) were recruited. All participants were screened for diabetes, neurological disorders, MRI compatibility, and pregnancy (for females). They followed a high-protein, low-sugar diet, fasted for six hours, and had their blood sugar levels checked before the scan.

Resting-state data were collected using the PMEEN system, including FDG-PET, fMRI, EEG, fNIRS, and eye tracking. Participants received an FDG dose based on body weight, and data acquisition began 30 minutes post-injection. The imaging parameters were optimized for PET, fMRI, and structural MRI sequences. Preprocessing for each modality followed standard procedures, with specific adjustments for resting-state fMRI. PET hypometabolism was quantified using NeuroQ, and functional connectivity maps were computed for fMRI, fNIRS, and EEG data. Functional connectivity strength was assessed through cross-correlation, and pupil size data were processed as described in previous sections. The detailed method is available in the Supplementary Method.

## Data and Code Availability

Data and materials for the experiment reported here are available upon request.

## Acknowledgments

This work was funded by the National Natural Science Foundation of China (grant no. 82027804 to Sijin Li).

## Author Contributions

Z.W., Y.G., S. Liu, L.H., X.S., and S. Li. designed the research. Z.W., Y.G., L.H., M.L., H.S. S.Y., J.Y., X.L., R.Z., Y.H., X.S., R.X., and S. Li. performed experiments. Z.W., Y.G., L.H., Y.H., M.L., H.S., S.Y., X.S., R.X., and S. Li. analyzed the data. Z.W., Y.G., L.H., X.S., and S. Li. wrote the manuscript. All authors edited the manuscript. Z.W., Y.G., J.C., S. Liu, L.H., and Y.L. contributed equally to this work.

## Competing Interests Statement

Y.G., L.H., J.Y., Y.H., X.S., and R.X. are employees of United Imaging Healthcare that manufacture imaging devices.

