## Supplementary Materials for "Concurrent mapping of Electrical, Chemical, and Functional Neuroactivity"

Shanxi Medical University

85 Jiefang South Road, Taiyuan, 030001, Shanxi, China

### Supplementary Method

### Acquistion of Healthy Volunteer Data with Visual Stimulation

Two participants were recruited for this experiment. A 46 to 50-year-old female participated in the task using the PMEEN system, while a 21 to 25-year-old male was recruited for the EEN experiment conducted in the lab. Two participants underwent screening for diabetes, personal or family history of neurological or neurodegenerative disorders, claustrophobia, MRI compatibility, and nuclear medicine safety. Female participants were additionally screened for current or potential pregnancy. Prior to the scan, participants were instructed to follow a high-protein, low-sugar diet for 24 hours, fast for six hours, and drink 2–6 glasses of water. Blood sugar levels were assessed prior to the experiments.

We used visual checkerboard stimulation to verify the data quality of each modality collected simultaneously in the PMEEN system. To ensure that each modality successfully captures the effects of visual stimulation, we adapted the experimental paradigms of the checkerboard task in FDG-PET and fMRI from previous studies^1,2^, as shown in Fig. S7. Specifically, an ‘embedded’ block design was performed to produce signal contrast for fast BOLD-fMRI, fNIRS, and EEG, as well as for the slower FDG-PET measurements. We compared pupil data and gaze position collected via the eye tracking system between the task-on and task-off conditions. The slow task/rest alternation periods were designed to generate FDG-PET contrast by alternating between task and rest over several minutes, while faster task on/off alternation periods were embedded within the stimulation to create contrast for BOLD-fMRI, fNIRS, and EEG.

We used the static FDG-PET to collect PET signals. Following a 30-minute setup to ensure optimal EEG impedance and fNIRS signal quality, FDG infusion was administered via the left cannula. Participants then rested for 5 minutes to achieve a calm and relaxed state. Afterward, we conducted a 25-minute simultaneous PMEEN session. Participants were positioned supine in the scanner bore with their heads placed in a 24-channel radiofrequency (RF) head-neck coil. Visual stimuli were presented using an infrared hot mirror mounted on the RF coil, reflecting an MR-compatible LCD screen. The infrared hot mirror was used to increase the accuracy of the eye tracking.

During the 25-minute checkerboard task, two 10 min visual stimulation periods were presented in an embedded 32/16 sec on/off design. The first 120 sec of the block was a sustained ‘on’ period to allow FDG-PET signal to rise from resting levels. During the ‘on’ periods (i.e., 120 s & 32 s periods), the visual stimulus was a circular checkerboard (size 39 cm, visual angle 9°), presented on a gray background. The checkerboard flickered (i.e., alternated black and white fields), at rate of 8 Hz. During the 16 s ‘off’ periods, participants rested with eyes open while viewing a white fixation cross (size 3 cm, visual angle 0°45′) presented on a gray background. Following the first 10 min stimulation block, a 3 min rest period followed, then the other 10 min stimulation. Participants were asked to try to keep their eyes open during the task.

FDG-PET, fMRI, EEG, eye-tracking, and fNIRS data were simultaneously collected during a 25-minute task window using the PMEEN systems. Participants received an injected dose of 18F-FDG at 7 mCi. During the task, PET data were acquired and reconstructed using the TOF approach with the following reconstruction parameters: 4 iterations, 20 subsets, Gaussian filter = 3 mm, matrix size = 512 × 512, 160 slices, slice thickness = 2 mm, field of view (FOV) = 300 mm × 300 mm, and voxel size = 1.6 mm × 1.6 mm × 2 mm. The PET data acquisition began with multiecho water-fat imaging (WFI) (scanning time = 30 seconds) and ultrashort TE (UTE) (scanning time = 5 minutes) MR sequences for PET attenuation correction. Following this, T2*-weighted echo planar imaging (EPI) signals were acquired over the 25-minute task window, simultaneously with the other imaging modalities, for further analysis in this study. The EPI parameters were as follows: TR = 2000 ms, TE = 30 ms, flip angle = 90°, slice thickness = 3.5 mm, 35 slices, FOV = 384 mm, and an in-plane resolution of 3.5 mm × 3.5 mm. After completing the task, T1-weighted images were acquired with the following parameters: TR = 7900 ms, TE = 3 ms, flip angle = 10°, FOV = 256 mm × 240 mm, slice thickness = 1 mm, 160 slices, and voxel size = 1 mm × 1 mm × 1 mm.

To examine the signal quality of EEG, fNIRS, and eye-tracking data collected by the PMEEN system, we set up the in-lab stimulation and collected the EEG, fNIRS, and eye-tracking data without the artifacts induced by PET or MR. This in-lab setup mirrored the conditions used in the simultaneous PMEEN acquisition but without the PET or MR artifacts. However, there were a few differences: the stimulus was presented on a stimulus computer, and participants viewed the checkerboard while seated in a chair.

To examine the fMRI signal quality collected by the PMEEN system, we collected only fMRI data during the same visual checkerboard task setup as the simultaneously PMEEN acquisition and compared it with the fMRI signal obtained from the PMEEN system.

The PMEEN data were preprocessed as below. The PET images were first temporally aligned with the fMRI signal and then the PET image was reconstructed with the parameters described above. Since fNIRS are collected simultaneously, attenuation correction of the fNIRS optic fibers based on the UTE and WFI images was then performed.

We preprocessed the rsfMRI BOLD data following a previous study^1^ using scripts from the 1000 Functional Connectomes Project^3^, with minor modifications. These scripts utilized FSL^4^ and AFNI^5^. We performed motion correction and skull stripping. The rsfMRI BOLD data were then spatially smoothed (FWHM = 4 mm). Both the anatomical T1-weighted images and rsfMRI BOLD data were registered to MNI space. Lastly, the rsfMRI BOLD data were high-pass filtered at a frequency of 0.01 Hz.

The EEG data were preprocessed to remove gradient and ballistocardiogram artifacts following a published algorithm^6^. We first aligned the time of EEG signals with functional MRI data by excluding EEG signals without MR gradient system trigger. Next, components identified as gradient artifacts were removed for each channel using singular value decomposition (SVD)^7^. The EEG data were then low-pass filtered (<125 Hz) and down-sampled to 250 Hz. Pulse artifacts were eliminated using independent component analysis (ICA), with components classified as artifactual based on their high similarity to the ECG signal. Data epochs containing significant artifacts were checked, but none were found. We then used EEGLAB^8^ to remove components with artifacts related to muscle, eye blink or eye movements. For the EEG data collected in lab, we ignored the step of gradient removal and others kept the same.

For the pupil size data, we first aligned the time of pupil size and gaze position data with functional MRI data by excluding signals without MR gradient system trigger. The blink artifacts were removed, and the pupil size time series was linearly detrended within each scan. We then calculated the pupil size amplitude by computing the root-mean-square envelope of the pupil size. Finally, the pupil size data were normalized to a range between 0 and 1, with the maximum value in each scanning run scaled to 1. For the eye tracking data collected in lab, the preprocessing steps were the same.

The fNIRS data were first screened manually to check for detector saturation, which occurred in none of the participants and none of the channels. Data epochs were checked for significant artifacts, but none were detected. After automatic detection (peak-to-peak >6 S.D.) and correction of spike ‘jumps’ using linear interpolation, optical intensity data were band-pass filtered between 0.01–0.1Hz, followed by converting into optical density variations (ΔOD). ΔOD timeseries for both wavelengths of interest were then transformed into relative concentration changes of oxyhemoglobin (Δ[HbO]) and deoxyhemoglobin (Δ[HbR]), respectively. For the fNIRS data collected in lab, the preprocessing steps were the same.

Since only static PET was collected, no further analysis after preprocessing steps was conducted. For the fMRI and fNIRS data, we averaged the signals separately during task-on and task-off periods to assess visual activation during the checkerboard task. The fMRI or fNIRS task-on map was then averaged over time points under task-on conditions. The time course of fMRI activation during task-on conditions was calculated by averaging the voxels within the visual cortex, as defined by the Hammers atlas^9,10^. Similarly, the time course of fNIRS activation was calculated by averaging the channels within the visual cortex during task-on conditions, specifically channels 13-15 and 28-30. Additionally, pupil size and gaze position were compared between task-on and task-off conditions.

To reduce artifacts, all the EEG data were first average re-referenced. For EEG spectrum analysis, we first segmented the EEG time series into task-on and task-off conditions. For each condition, we computed the power spectrum using a multi-taper time-frequency transformation (with a 1 s window, 0.05 s step, 10 tapers, 250 Hz sampling rate, 1-30 Hz frequency range, and a padding factor of 5), implemented via Chronux11, for the three visual cortex channels (O1, O2, and Oz), followed by calculating the mean across channels and conditions. The resulting mean power spectrum was then converted to decibel units. The difference in spectra between task-on and task-off conditions was computed by subtracting the task-off spectrum from the task-on spectrum and averaging the resulting spectra over conditions. To estimate EEG power time course at the fundamental frequency (8 Hz) and harmonic frequencies (16 Hz and 24 Hz) of the checkerboard stimulation during task-on conditions averaged over the three visual cortex channels (O1, O2, and Oz), we first computed the spectrogram for each EEG channel using a multi-taper time-frequency transformation (with a 1 s window, 0.02 s step, 3 tapers, 250 Hz sampling rate, 1-30 Hz frequency range, and a padding factor of 5), implemented via Chronux11. The EEG power time course at specific frequency bands (8Hz, 16Hz ,24Hz) was obtained by averaging the spectrogram within the specific frequency bins across all three channels (O1, O2, and Oz) under task-on conditions, followed by converted to decibel units and normalized between 0 and 1.

The initial two 120-second intervals of the two 10-minute checkerboard tasks were excluded from the analysis, as their purpose was to allow FDG-PET signal to rise from resting levels, and this duration was longer compared to other segments.

### Acquisition of Patient Resting-State Data

One healthy participant (female, age 41 to 45) was recruited as a control for this experiment. Additionally, three participants with specific conditions were recruited: a 36 to 40-year-old male with major depressive disorder, a 56 to 60-year-old female with Alzheimer's disease (AD), and a 26 to 30-year-old female with epilepsy. All participants were asked about their history of diabetes, personal or family history of neurological or neurodegenerative disorders, claustrophobia, MRI compatibility, and nuclear medicine safety. Female participants were additionally screened for current or potential pregnancy. Prior to the scan, all the participants were instructed to follow a high-protein, low-sugar diet for 24 hours, fast for six hours, and drink 2–6 glasses of water. Blood sugar levels were assessed prior to the experiments.

Participants were imaged in a resting state using the simultaneous PMEEN system. They were instructed to keep their eyes open and remain relaxed during the scan. Static FDG-PET data were collected, and to optimize the signal-to-noise ratio, PMEEN imaging was initiated 30 minutes post-injection of 18F-FDG, when the tracer had reached uniform distribution across the brain, minimizing blood flow-dependent variations^12^. Since it was a resting-state session, we only recorded pupil size for the eye tracking during the scan.

FDG-PET, fMRI, EEG, eye-tracking, and fNIRS data were simultaneously collected during a 20-minute resting-state window using the PMEEN systems after injection. The participants received an injected dose of 4.44 MBq per kilogram of body weight. PET data were acquired and reconstructed using the TOF approach with the following reconstruction parameters: 4 iterations, 20 subsets, Gaussian filter = 3 mm, matrix size = 512 × 512, 160 slices, slice thickness = 2 mm, field of view (FOV) = 300 mm × 300 mm, and voxel size = 1.6 mm × 1.6 mm × 2 mm. PET data acquisition began with multiecho water-fat imaging (WFI) (scanning time = 30 seconds) and ultrashort TE (UTE) (scanning time = 5 minutes) MR sequences for PET attenuation correction. Following this, T2*-weighted echo planar imaging (EPI) signals were acquired over 6.3 minutes, simultaneously with the other imaging modalities, for further analysis in this study. The EPI parameters were as follows: TR = 2100 ms, TE = 30 ms, flip angle = 90°, slice thickness = 3.5 mm, 35 slices, FOV = 384 mm, and in-plane resolution = 3.5 mm × 3.5 mm. Afterward, T1-weighted images were acquired with the following parameters: TR = 7900 ms, TE = 3 ms, flip angle = 10°, FOV = 256 mm × 240 mm, slice thickness = 1 mm, 160 slices, and voxel size = 1 mm × 1 mm × 1 mm. Finally, additional MR structural sequences were obtained, including T2-FLAIR, QSM, ASL, and DTI.

### The preprocessing for each modality, except for fMRI where we made some modifications for the resting-state data, followed the same procedures as those used for the visual checkerboard task, as detailed in the section of “Acquistion of Healthy Volunteer Data with Visual Stimulation”. For the resting-state fMRI data, motion correction and skull stripping were performed. The data were then spatially smoothed with a full width at half maximum (FWHM) of 4 mm. Subsequently, both the anatomical images and rsfMRI BOLD data were registered to MNI space, and nuisance parameters were regressed out, including linear and quadratic trends, motion parameters, white matter, global and CSF signals. Finally, the rsfMRI BOLD data were temporally smoothed within the 0.01 – 0.1 Hz frequency band.

The hypometabolism in the PET images of each patient was quantified using NeuroQ (version 3.80, Syntermed, Inc., Atlanta, GA, United States). NeuroQ is a commercial neurological application that provides a comprehensive analysis of 240 brain regions, comparing the patient's scan to those from an asymptomatic control group, and offering a schematic summary of the results.

A correlation map for a seed region was computed for each fMRI session. The time course for the seed region was derived by averaging the time courses of all the voxels within that seed region. Seed regions were defined based on brain atlas from the Hammers atlas^9,10^. This time course served as the reference to compute functional connectivity for the seed region. The strength of functional connectivity was quantified as the cross-correlation coefficient between the reference time course and the time course of each voxel in the brain. We then applied a threshold, retaining only correlations with an absolute value greater than a specific value and clusters larger than 200 voxels.

Similarly, a correlation map for a seed region was generated for each fNIRS session. The time course for the seed region was derived by averaging the time courses of all the channels within that region. Seed regions were defined based on the MNI position of the fNRIS channels (Table S3). This time course served as the reference to compute functional connectivity for the seed region. The strength of functional connectivity was quantified as the cross-correlation coefficient between the reference time course and the time course of each channel in the fNIRS signal. We then applied a threshold, retaining only correlations with an absolute value greater than a specific value. The fNIRS functional connectivity matrix for each session was computed by cross correlating the time courses of all 30 fNIRS channels using Pearson’s correlation coefficient.

For EEG data, we computed the spectrogram for each channel using a multi-taper time-frequency transformation (1 s window, 0.02 s step, 3 tapers, 250 Hz sampling rate, 1-30 Hz frequency range, padding factor of 5), implemented with Chronux^11^. The mean spectrogram for specific EEG channels was obtained by averaging the spectrograms across these channels. The resulting mean power spectrogram was then converted to decibel units and normalized at each frequency bin by subtracting the mean and dividing by the standard deviation. To compute a correlation map for an EEG frequency band, we first extracted the time courses for different frequency bands across all channels. The spectrogram for each channel was calculated using a multi-taper time-frequency transformation (1 s window, 0.02 s step, 3 tapers, 250 Hz sampling rate, 1-30 Hz frequency range, padding factor of 5), implemented with Chronux^11^, and normalized by subtracting the mean and dividing by the standard deviation. The normalized spectrogram was then averaged within the delta (0.5–4 Hz), theta (4–8 Hz), alpha (8–12 Hz), and beta (12–30 Hz) frequency bands. For each frequency band, the correlation map for each seed channel was generated by calculating the cross-correlation coefficient between the time course of the frequency band from the seed channel and that from each other channel. We then applied a threshold, retaining only correlations with an absolute value greater than a specific value.

### Pupil size was processed as described in section of “Acquistion of Healthy Volunteer Data with Visual Stimulation”.

### Supplementary Results

### Clinical Application

**AD patient**

The detailed results of BOLD-fMRI correlation maps in the AD patient were shown below (Fig. 5B). In the left frontal seed map, the AD patient exhibited positive correlations in the superior frontal gyrus, middle frontal gyrus, angular gyrus, lateral occipital cortex (superior division), cingulate gyrus, precuneus cortex, paracingulate gyrus, frontal pole, inferior frontal gyrus, middle temporal gyrus (posterior division), and left thalamus. Negative correlations were observed in the lateral occipital cortex, precuneus cortex, superior parietal lobule, occipital pole, occipital fusiform gyrus, and lingual gyrus, whereas the healthy control displayed broader and larger positive correlations in the middle frontal gyrus, superior frontal gyrus, paracingulate gyrus, frontal medial cortex, precuneus cortex, temporal pole, frontal pole, left accumbens, and superior temporal gyrus (anterior division), and negative correlations in the precentral gyrus, lateral occipital cortex (superior division), superior parietal lobule, angular gyrus, supramarginal gyrus (anterior division), frontal pole, and cerebellum (Fig. 5B). In the right frontal seed map, the AD patient showed positive correlation in the superior frontal gyrus, angular gyrus, frontal pole, middle frontal gyrus, paracingulate gyrus, and negative correlation in the lateral occipital cortex, cuneal cortex, middle temporal gyrus (posterior division), supramarginal gyrus (posterior division), planum temporale, precuneus cortex, while the healthy control showed positive correlations in precentral gyrus, superior frontal gyrus, precuneus cortex, supramarginal gyrus (posterior division), lateral occipital cortex (superior division), paracingulate gyrus, angular gyrus, inferior frontal gyrus (pars opercularis), frontal lobe, left thalamus, brain stem, with negative correlations in the insular cortex, lateral occipital cortex (superior division), cingulate gyrus (posterior division), lingual gyrus (Fig. 5B).

For the left parietal seed map, the AD patient exhibited positive correlations in the postcentral gyrus, precentral gyrus, lateral occipital cortex (superior division), supplementary motor cortex, superior frontal gyrus, cingulate gyrus (anterior division), paracingulate gyrus, supramarginal gyrus, planum polare, central opercular cortex, and frontal operculum cortex. The AD patient showed negative correlations in the precuneus cortex, lingual gyrus, and occipital pole. In contrast, the healthy control demonstrated positive correlations in the superior frontal gyrus, superior parietal lobule, precentral gyrus, supplementary motor cortex, postcentral gyrus, and anterior division of the supramarginal gyrus (Fig. 5B), with negative correlations in the frontal orbital cortex and left thalamus. In the right parietal seed map, the AD patient showed positive correlations in the precentral gyrus, superior parietal lobule, superior frontal gyrus, postcentral gyrus, lateral occipital cortex (superior division), middle frontal gyrus, precuneus cortex, supramarginal gyrus, angular gyrus, parietal operculum cortex, and central opercular cortex, with negative correlations in the cuneal cortex and lingual gyrus. In contrast, the healthy control exhibited positive correlations in the superior parietal lobule, postcentral gyrus, superior frontal gyrus, supplementary motor cortex, precentral gyrus, middle frontal gyrus, paracingulate gyrus, angular gyrus, supramarginal gyrus (anterior division), and brainstem, with negative correlations in the cingulate gyrus (posterior division) and the occipital pole (Fig. 5B). In the cingulate cortex seed map, the AD patient showed positive correlations in the supplementary motor cortex, superior frontal gyrus, angular gyrus, middle frontal gyrus, paracingulate gyrus, cingulate gyrus, frontal pole, precuneus cortex, central opercular cortex, middle temporal gyrus (posterior division), lateral occipital cortex (superior division), left thalamus, and brainstem, with negative correlations in the lateral occipital cortex, cuneal cortex, and middle temporal gyrus (temporooccipital part). The healthy control displayed positive correlations in the superior frontal gyrus, middle frontal gyrus, cingulate gyrus, frontal pole, precuneus cortex, paracingulate gyrus, lateral occipital cortex (superior division), and planum temporale, with negative correlations in the superior division of the lateral occipital cortex, superior parietal lobule, and supramarginal gyrus (posterior division) (Fig. 5B).

**Epilepsy patient**

The specific results of BOLD-fMRI correlation maps in the epilepsy patient using the left frontal lobe as the seed region were described below. The epilepsy patient exhibited positive correlations in the central opercular cortex, middle temporal gyrus (temporooccipital part), angular gyrus, parietal operculum cortex, insular cortex, inferior frontal gyrus (pars opercularis), frontal orbital cortex, frontal pole, cuneal cortex, subcallosal cortex, cingulate gyrus (posterior division), temporal pole, brainstem, and cerebellum. Negative correlations were observed in the inferior temporal gyrus (temporooccipital part), precentral gyrus, superior parietal lobule, lateral occipital cortex (superior division), postcentral gyrus, superior frontal gyrus, brainstem, and cerebellum. In contrast, the healthy control showed broader and stronger positive correlations in the middle frontal gyrus, superior frontal gyrus, paracingulate gyrus, frontal medial cortex, precuneus cortex, temporal pole, frontal pole, left accumbens, and superior temporal gyrus (anterior division), with negative correlations in the precentral gyrus, lateral occipital cortex (superior division), superior parietal lobule, angular gyrus, supramarginal gyrus (anterior division), frontal pole, and cerebellum (Fig. 6C). The fNIRS HbO-based functional connectivity matrix revealed that the epilepsy patient had increased connectivity within the right parietal lobe, but decreased connectivity in other regions, relative to the healthy control (Fig. 6D). Similarly, using the left frontal lobe as a seed region, the correlation map calculated using fNIRS HbO signal showed reduced connectivity (values >0.5) in the epilepsy patient compared to the healthy control (Fig. 6D).

### Supplementary Figures

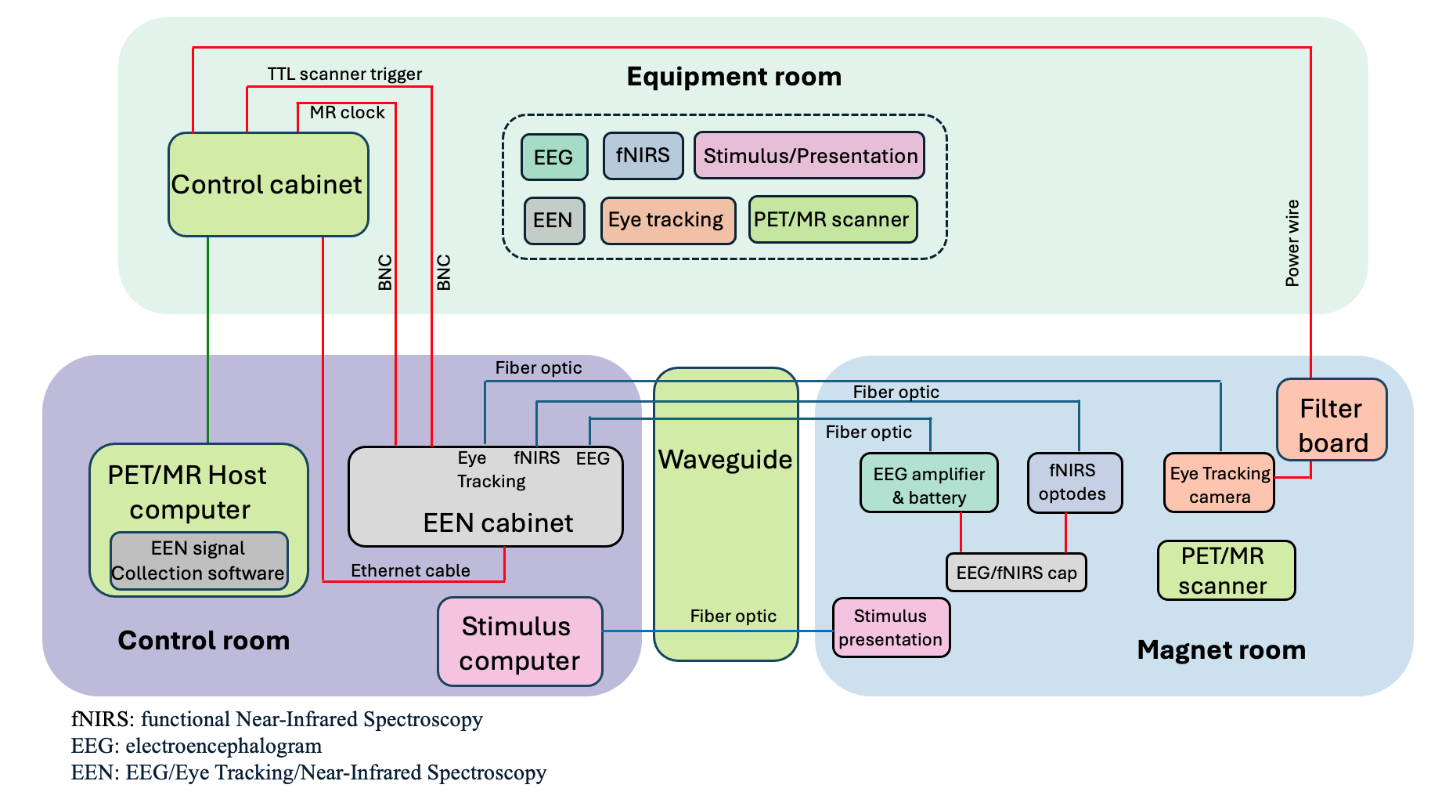

Figure S1. The schematic of the PMEEN setup.

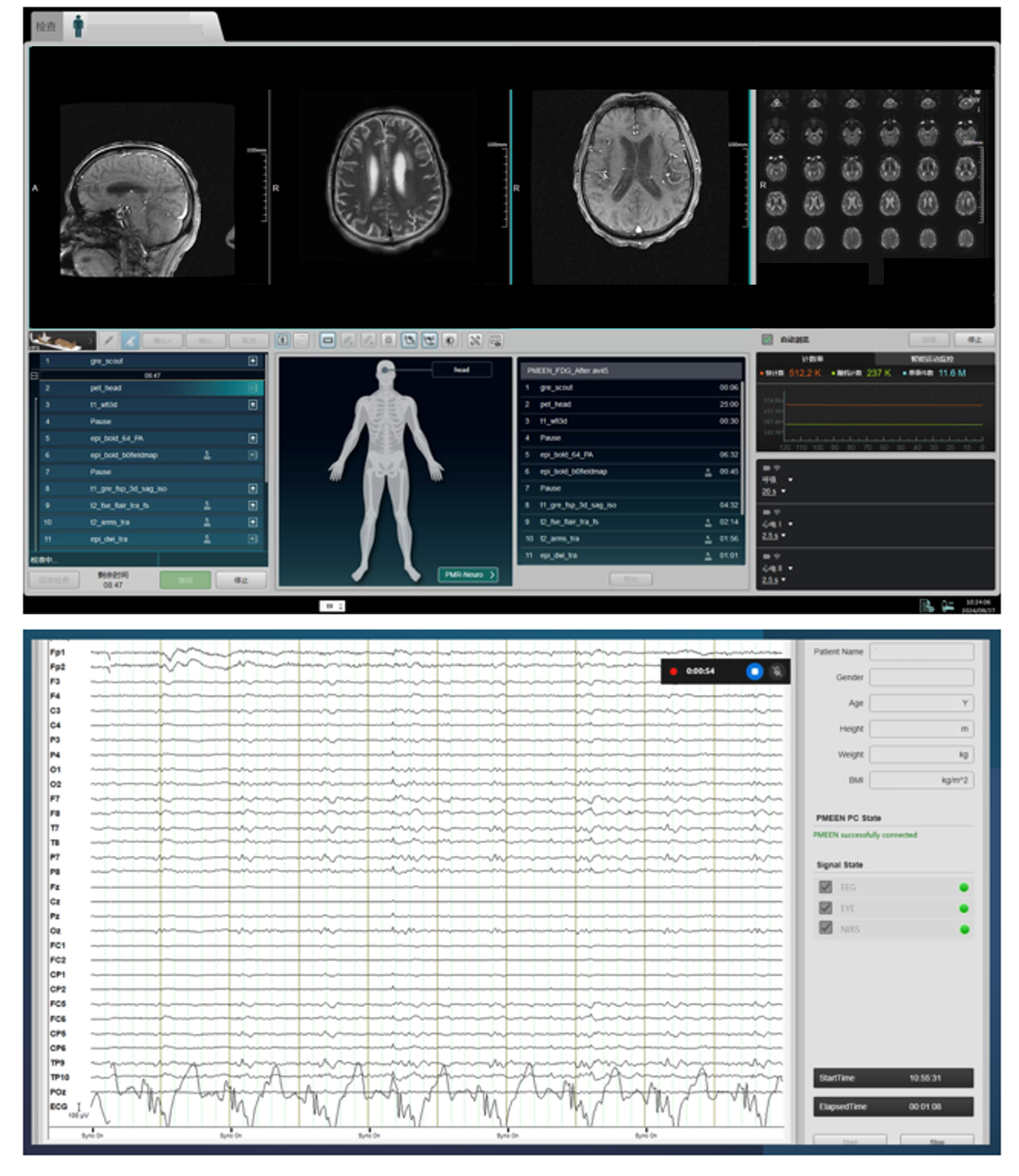

Figure S2. The PMEEN software for signal collection. Top: the example of PET and MR signal collection. Bottom: the example of fNIRS, EEG and eye tracking signal collection.

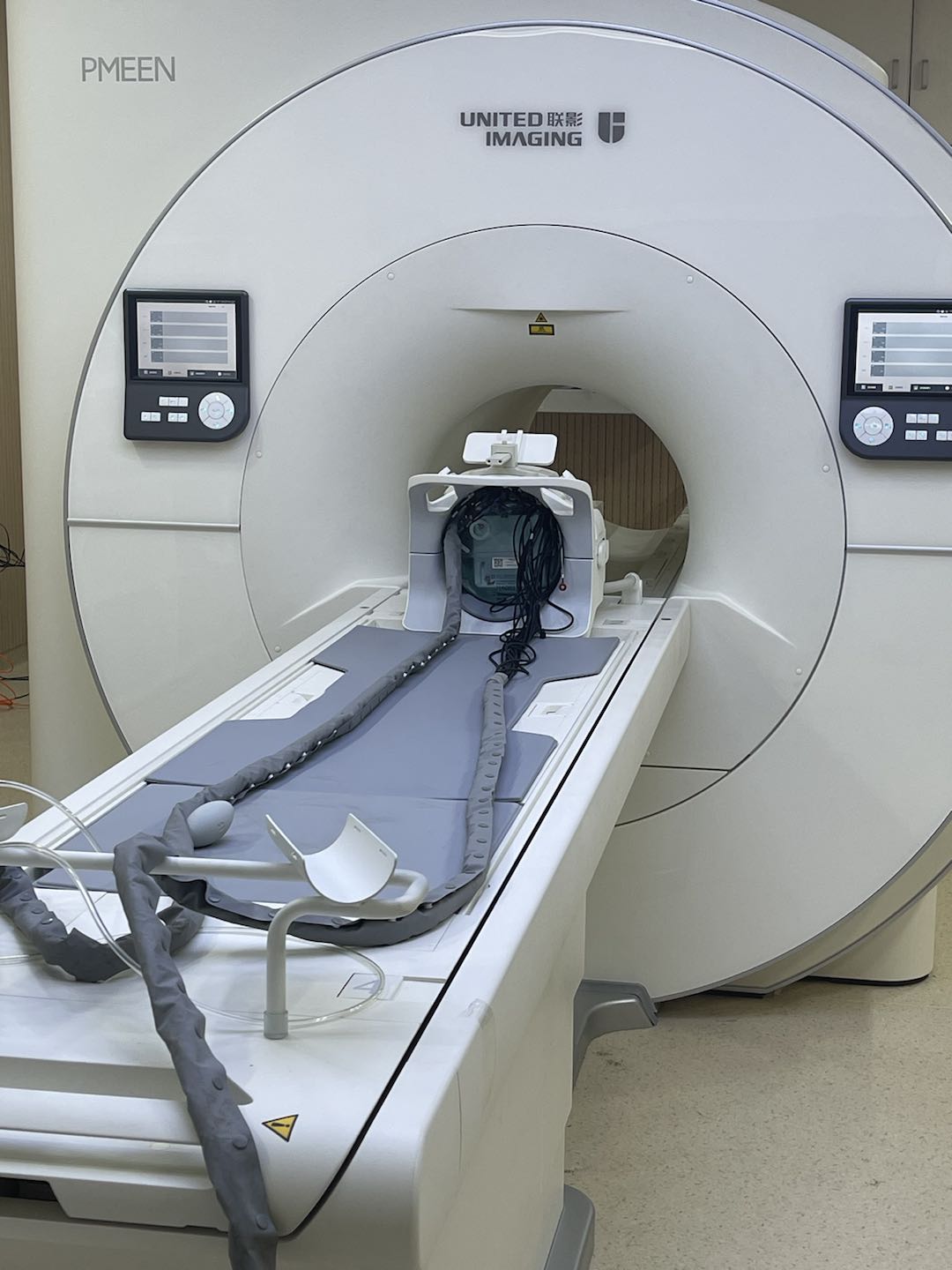

Figure S3. Experimental setup for evaluating the impact of the EEG electrodes, Eye-tracking mirror, fNIRS optodes and optic fiber on MR images, as well as the MR images acquired from the PMEEN system.

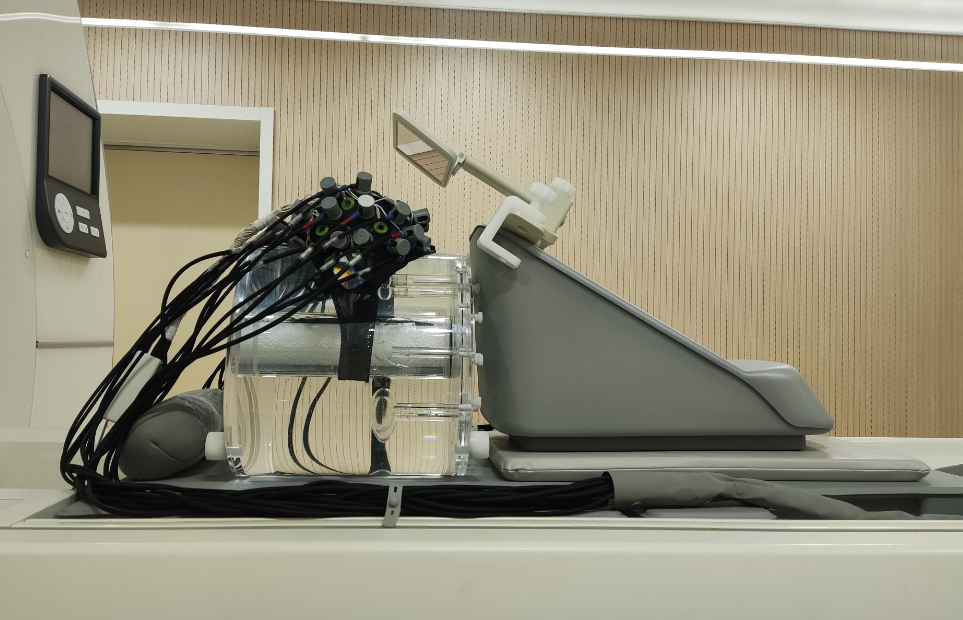

Figure S4. Experimental setup for evaluating the impact of the EEG/fNIRS cap, fNIRS optodes, optical fibers, and eye-tracking mirror on PET IQ images acquired from the PMEEN system.

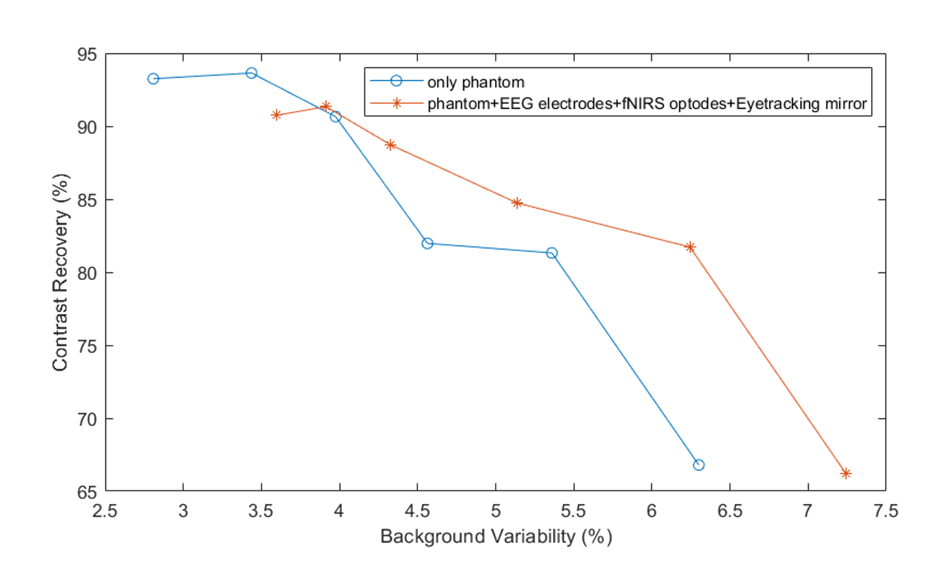

Figure S5. Contrast recovery vs. background variability for each sphere in the PET IQ phantom (with MR pulsing) was reconstructed using OSEM with 1 iteration. Two conditions were tested: the first with the phantom only, and the second with the phantom fitted with the EEG/fNIRS cap, fNIRS optodes, optical fibers, and eye-tracking mirror.

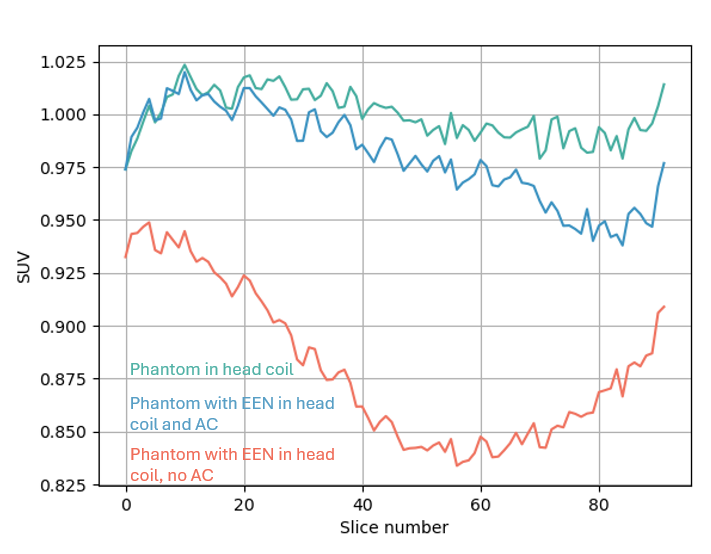

Figure S6. The changes in PET SUV of the PMEEN system induced by **E**EG electrodes, **E**ye-tracking mirror, f**N**IRS optodes and optical fiber (EEN), with (blue) or without (orange) attenuation correction (AC) for EEN components, compared to the condition with only the uniform cylindrical phantom inside the head coil (green).

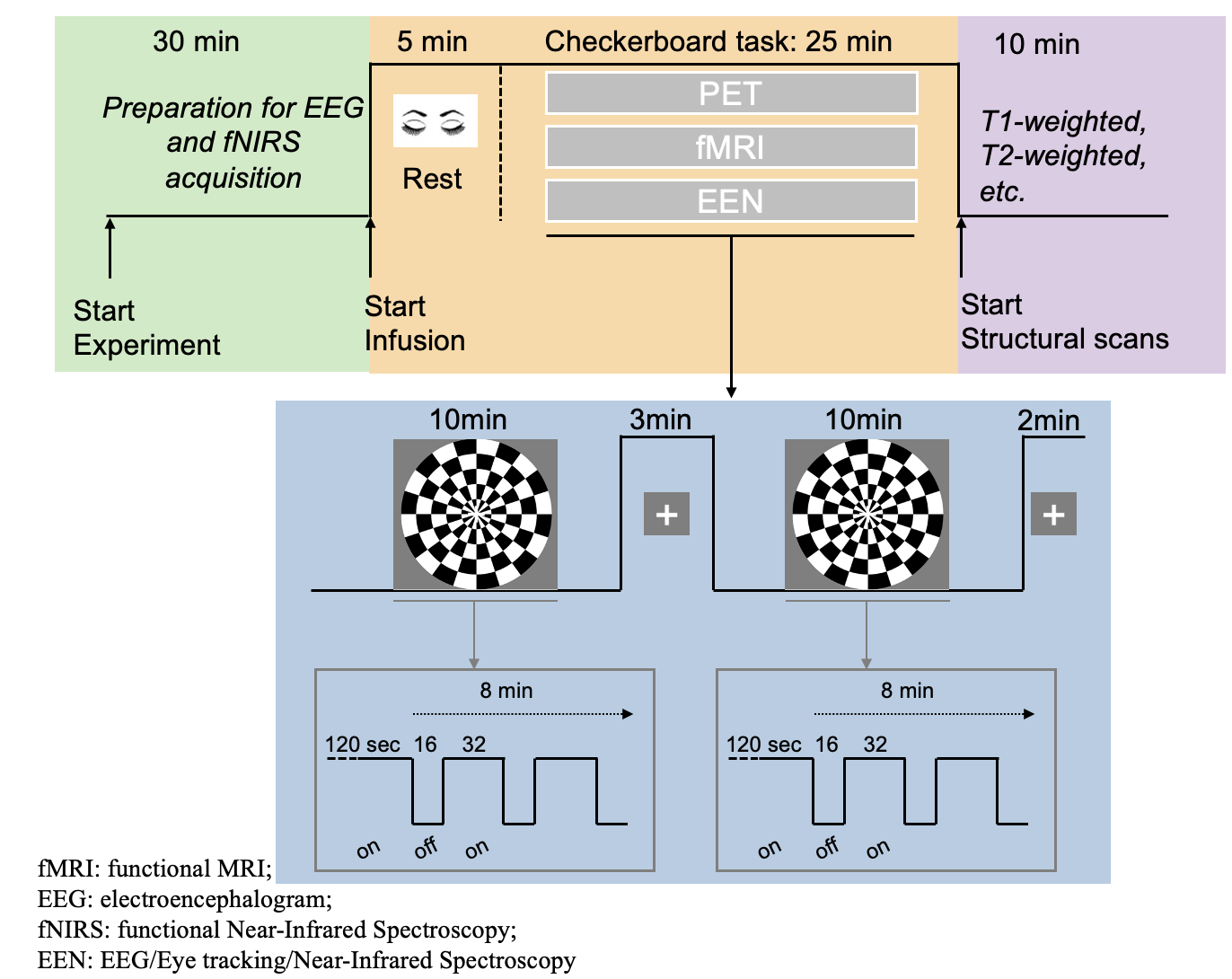

Figure S7. The experimental protocol of checkerboard task.

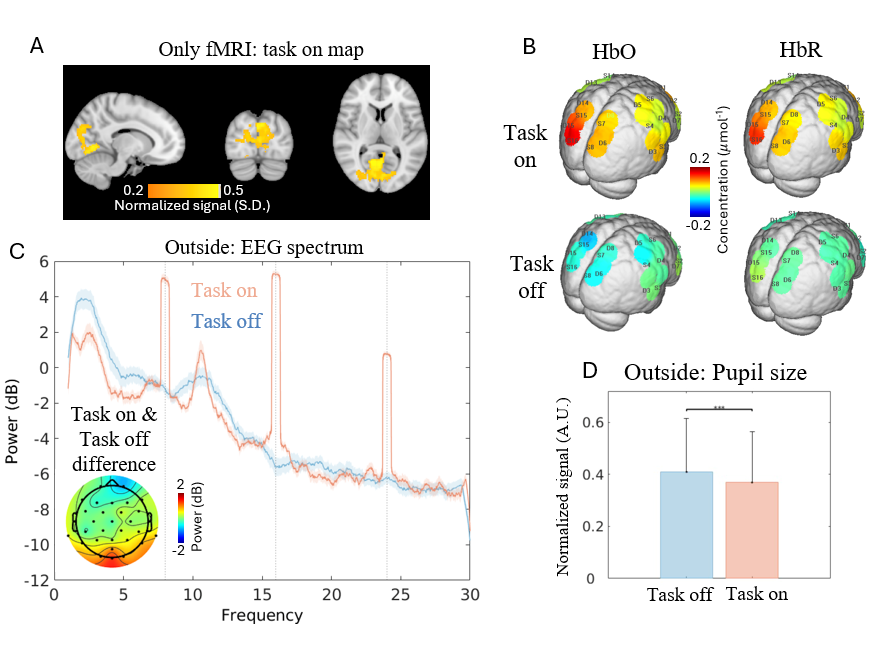

Figure S8. (A). The fMRI was collected without other imaging modalities while the participant performed the visual checkerboard task. (B-D). The results from visual checkerboard task collected outside the PMEEN scanner.

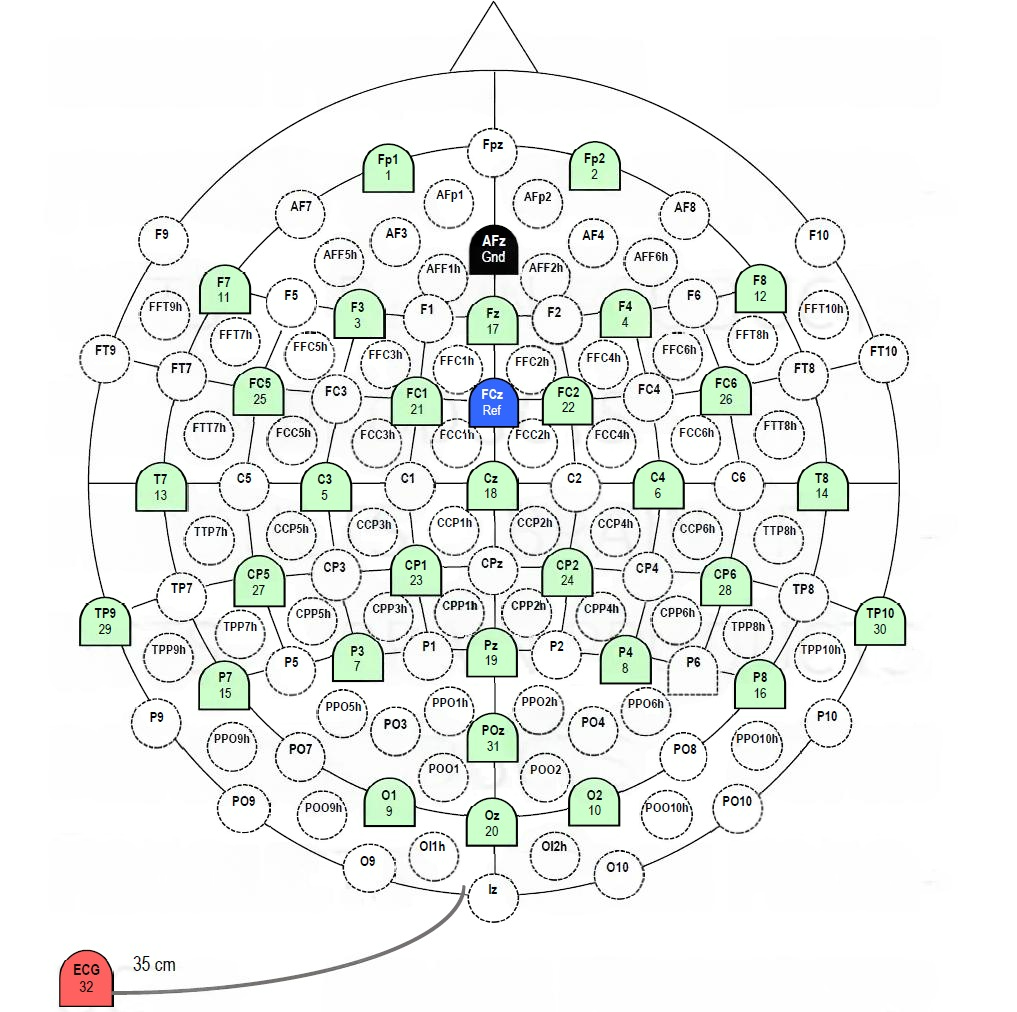

Figure S9. The colored green channels represent the 31 EEG channel location used in PMEEN system, adapted from Brain Products. Red: ECG channel; Black: ground channel; Blue: reference channel.

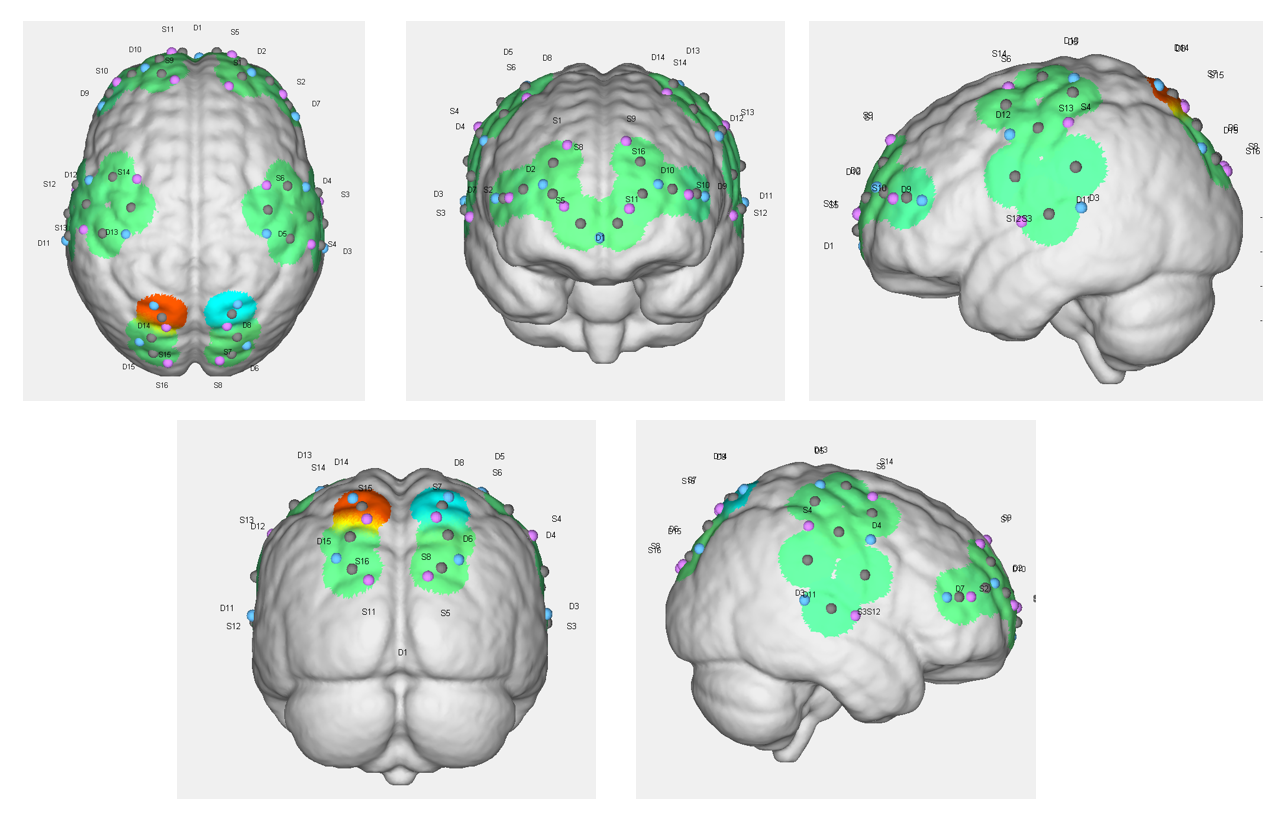

Figure S10. The location of the fNIRS sources and detectors on the brain.

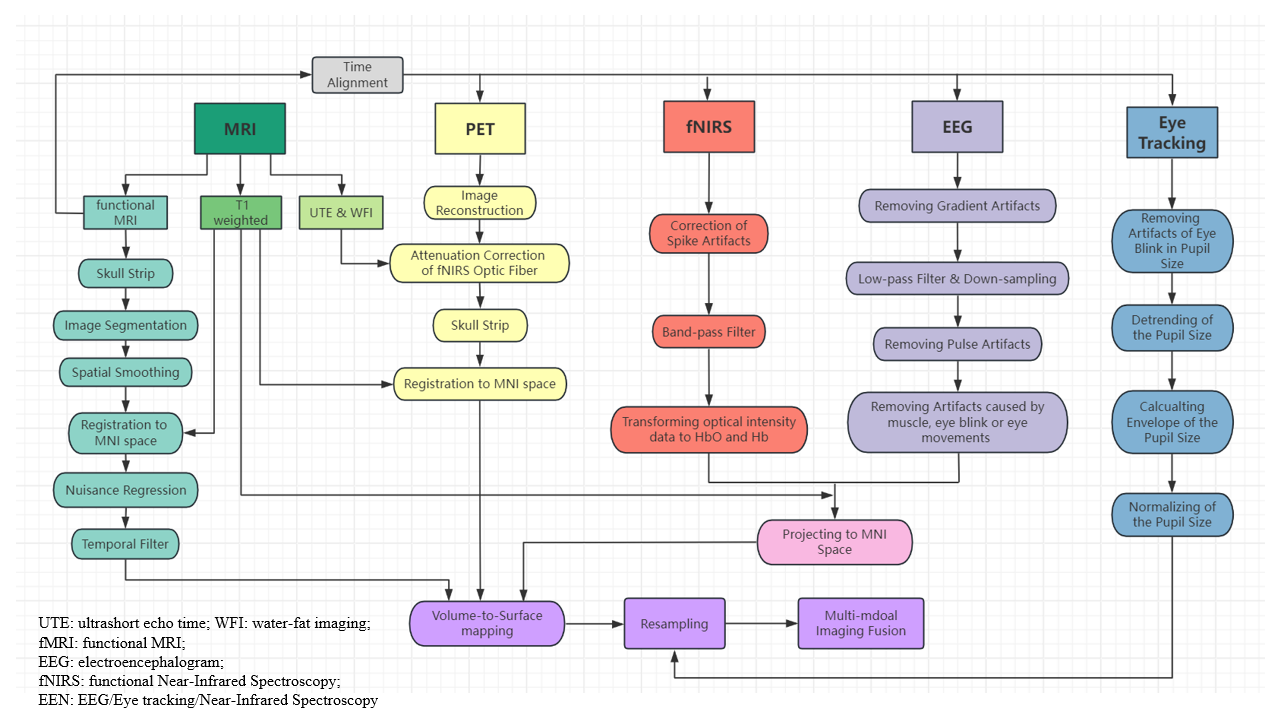

Figure S11. The preprocessing of each imaging modality in the PMEEN system and the multi-modal imaging fusion.

### Supplementary Tables

Table S1. MR ACR results derived with MR only and with the PET sequence on, EEG/fNIRS cap, fNIRS optodes and optic fiber, and eye tracking mirror positioned on the phantom respectively.

|  | Only MRI | PMEEN |
| --- | --- | --- |
| Uniformity (%) | 92 | 91 |
| Slice position accuracy (mm) | 1.7 | 3.7 |
| Spatial resolution (mm) | 1 | 1 |
| Slice thickness accuracy (mm) | 0.12 | 0.17 |
| Percent signal ghosting (%) | 0.10 | 0.32 |
| Signal-to-noise ratio | 1555.4 | 1553.4 |
| Conclusion | Pass | Pass |

Table S2. NEMA IQ results obtained with the MR sequence (BOLD EPI) on, EEG/fNIRS cap, and eye tracking mirror positioned on the phantom.

| Sphere diameter (mm) | 10 | 13 | 17 | 22 | 28 | 37 |
| --- | --- | --- | --- | --- | --- | --- |
| Hot contrast (%) | 66.22 | 81.72 | 84.73 | 88.71 | 91.34 | 90.75 |
| Hot contrast acceptable range (%) | ≥45 | ≥60 | ≥65 | ≥65 | ≥65 | ≥70 |
| Background variability (%) | 7.24 | 6.24 | 5.14 | 4.33 | 3.91 | 3.60 |
| Background variability acceptable range (%) | <9 | <8 | <7 | <7 | <7 | <7 |
| Lung residual error (%) | 3.7 | | | | | |
| Lung residual error acceptable range (%) | ≤10 | | | | | |
| Conclusion | Pass | | | | | |

Table S3. The MNI position and Brodmann Area of fNIRS channels.

| Channel Number  (Source-Detector) | MNI Position | | | Brodmann Area | Percentage |
| --- | --- | --- | --- | --- | --- |
| CH1 (S1-D2) | 25.958 | 59.688 | 30.706 | 9 - Dorsolateral prefrontal cortex | 0.2603 |
|  |  |  |  | 10 - Frontopolar area | 0.2851 |
|  |  |  |  | 46 - Dorsolateral prefrontal cortex | 0.4545 |
| CH2 (S2-D2) | 41.262 | 57.222 | 17.224 | 10 - Frontopolar area | 0.2329 |
|  |  |  |  | 45 - pars triangularis Broca's area | 0.008 |
|  |  |  |  | 46 - Dorsolateral prefrontal cortex | 0.759 |
| CH3 (S2-D7) | 51.485 | 43.642 | 12.7 | 45 - pars triangularis Broca's area | 0.6679 |
|  |  |  |  | 46 - Dorsolateral prefrontal cortex | 0.3321 |
| CH4 (S3-D3) | 71.406 | -22.13 | 6.7696 | 21 - Middle Temporal gyrus | 0.295 |
|  |  |  |  | 22 - Superior Temporal Gyrus | 0.705 |
| CH5 (S3-D4) | 68.141 | -4.8864 | 24.103 | 4 - Primary Motor Cortex | 0.0094 |
|  |  |  |  | 6 - Pre-Motor and Supplementary Motor Cortex | 0.0346 |
|  |  |  |  | 22 - Superior Temporal Gyrus | 0.0126 |
|  |  |  |  | 43 - Subcentral area | 0.9434 |
| CH6 (S4-D3) | 69.776 | -34.25 | 31.804 | 2 - Primary Somatosensory Cortex | 0.288 |
|  |  |  |  | 22 - Superior Temporal Gyrus | 0.0095 |
|  |  |  |  | 40 - Supramarginal gyrus part of Wernicke's area | 0.538 |
|  |  |  |  | 48 - Retrosubicular area | 0.1646 |
| CH7 (S4-D4) | 63.424 | -18.177 | 46.203 | 1 - Primary Somatosensory Cortex | 0.5302 |
|  |  |  |  | 2 - Primary Somatosensory Cortex | 0.0107 |
|  |  |  |  | 3 - Primary Somatosensory Cortex | 0.2847 |
|  |  |  |  | 4 - Primary Motor Cortex | 0.1744 |
| CH8 (S4-D5) | 52.586 | -30.399 | 61.821 | 1 - Primary Somatosensory Cortex | 0.3954 |
|  |  |  |  | 2 - Primary Somatosensory Cortex | 0.1255 |
|  |  |  |  | 3 - Primary Somatosensory Cortex | 0.384 |
|  |  |  |  | 4 - Primary Motor Cortex | 0.0114 |
|  |  |  |  | 40 - Supramarginal gyrus part of Wernicke's area | 0.0837 |
| CH9 (S5-D1) | 12.004 | 73.37 | -0.2778 | 10 - Frontopolar area | 0.7111 |
|  |  |  |  | 11 - Orbitofrontal area | 0.2889 |
| CH10 (S5-D2) | 26.342 | 67.479 | 14.808 | 10 - Frontopolar area | 0.993 |
|  |  |  |  | 46 - Dorsolateral prefrontal cortex | 0.007 |
| CH11 (S6-D4) | 51.271 | -1.1162 | 55.627 | 4 - Primary Motor Cortex | 0.0837 |
|  |  |  |  | 6 - Pre-Motor and Supplementary Motor Cortex | 0.9163 |
| CH12 (S6-D5) | 40.408 | -14.524 | 69.666 | 4 - Primary Motor Cortex | 0.4291 |
|  |  |  |  | 6 - Pre-Motor and Supplementary Motor Cortex | 0.5709 |
| CH13 (S7-D6) | 23.329 | -85.091 | 49.443 | 7 - Somatosensory Association Cortex | 0.4375 |
|  |  |  |  | 19 - V3 | 0.5625 |
| CH14 (S7-D8) | 20.41 | -72.311 | 63.649 | 7 - Somatosensory Association Cortex | 1 |
| CH15 (S8-D6) | 20.162 | -94.662 | 33.559 | 18 - Visual Association Cortex (V2) | 0.6871 |
|  |  |  |  | 19 - V3 | 0.3129 |
| CH16 (S9-D10) | -18.704 | 61.14 | 31.276 | 9 - Dorsolateral prefrontal cortex | 0.3424 |
|  |  |  |  | 10 - Frontopolar area | 0.4436 |
|  |  |  |  | 46 - Dorsolateral prefrontal cortex | 0.214 |
| CH17 (S10-D9) | -47.481 | 50.39 | 14.981 | 45 - pars triangularis Broca's area | 0.369 |
|  |  |  |  | 46 - Dorsolateral prefrontal cortex | 0.631 |
| CH18 (S10-D10) | -35.186 | 60.754 | 17.385 | 10 - Frontopolar area | 0.4206 |
|  |  |  |  | 46 - Dorsolateral prefrontal cortex | 0.5794 |
| CH19 (S11-D1) | -7.1829 | 73.106 | -0.1086 | 10 - Frontopolar area | 0.7796 |
|  |  |  |  | 11 - Orbitofrontal area | 0.2204 |
| CH20 (S11-D10) | -19.732 | 69.209 | 14.86 | 10 - Frontopolar area | 1 |
| CH21 (S12-D11) | -70.593 | -16.446 | 7.2602 | 21 - Middle Temporal gyrus | 0.196 |
|  |  |  |  | 22 - Superior Temporal Gyrus | 0.804 |
| CH22 (S12-D12) | -65.927 | -0.48834 | 24.876 | 4 - Primary Motor Cortex | 0.0228 |
|  |  |  |  | 6 - Pre-Motor and Supplementary Motor Cortex | 0.2573 |
|  |  |  |  | 43 - Subcentral area | 0.7199 |
| CH23 (S13-D11) | -70.267 | -28.888 | 29.394 | 2 - Primary Somatosensory Cortex | 0.6556 |
|  |  |  |  | 22 - Superior Temporal Gyrus | 0.0132 |
|  |  |  |  | 40 - Supramarginal gyrus part of Wernicke's area | 0.1523 |
|  |  |  |  | 42 - Primary and Auditory Association Cortex | 0.0199 |
|  |  |  |  | 48 - Retrosubicular area | 0.1589 |
| CH24 (S13-D12) | -60.407 | -11.442 | 47.516 | 1 - Primary Somatosensory Cortex | 0.1296 |
|  |  |  |  | 3 - Primary Somatosensory Cortex | 0.3111 |
|  |  |  |  | 4 - Primary Motor Cortex | 0.3333 |
|  |  |  |  | 6 - Pre-Motor and Supplementary Motor Cortex | 0.1926 |
|  |  |  |  | 43 - Subcentral area | 0.0333 |
| CH25 (S13-D13) | -51.436 | -27.499 | 64.106 | 1 - Primary Somatosensory Cortex | 0.3633 |
|  |  |  |  | 2 - Primary Somatosensory Cortex | 0.0408 |
|  |  |  |  | 3 - Primary Somatosensory Cortex | 0.5143 |
|  |  |  |  | 4 - Primary Motor Cortex | 0.0816 |
| CH26 (S14-D12) | -45.709 | 3.7983 | 58.297 | 6 - Pre-Motor and Supplementary Motor Cortex | 0.9835 |
|  |  |  |  | 9 - Dorsolateral prefrontal cortex | 0.0165 |
| CH27 (S14-D13) | -35.607 | -13.354 | 71.338 | 4 - Primary Motor Cortex | 0.2612 |
|  |  |  |  | 6 - Pre-Motor and Supplementary Motor Cortex | 0.7388 |
| CH28 (S15-D14) | -18.534 | -74.273 | 63.192 | 7 - Somatosensory Association Cortex | 1 |
| CH29 (S15-D15) | -24.253 | -85.543 | 48.536 | 7 - Somatosensory Association Cortex | 0.4008 |
|  |  |  |  | 19 - V3 | 0.5992 |
| CH30 (S16-D15) | -23.296 | -94.055 | 32.925 | 18 - Visual Association Cortex (V2) | 0.6184 |
|  |  |  |  | 19 - V3 | 0.3816 |
